# Traumatic brain injury (TBI) and mortality in older adults with and without pre-injury dementia

**DOI:** 10.1101/2024.10.09.24314691

**Authors:** Helen Lai, Eyal Soreq, Niall J Bourke, Claire Baker, Karl A Zimmerman, Megan E Parkinson, Sarah JC Daniels, Edward W Gregg, David J Sharp, Lucia M Li

## Abstract

**Importance:** Incidence of traumatic brain injury (TBI) is rising in older adults. Dementia is a common comorbidity amongst older adults and may worsen post-TBI outcomes, but this has not been systematically studied.

**Objective:** To compare all-cause mortality following TBI or non-TBI trauma (NTT), and quantify the impacts of age, deprivation, and pre-injury dementia.

**Design:** Population-based retrospective cohort study

**Setting:** Electronic health records (EHRs) from primary and secondary care

**Participants:** Adult residents in Wales aged 18-100 with TBI or NTT from January 2000 to December 2022

**Exposures:** Traumatic (intracranial) brain injury (TBI) or non-TBI trauma (NTT) recorded in secondary care. Pre-injury diagnosis of dementia within primary or secondary care.

**Main Outcomes and Measures:** All-cause mortality within 1, 6, and 12 months of hospitalised TBI/NTT in those with and without pre-injury dementia. Cox proportional hazard models were used to estimate survival probability across groups. Groups were propensity-matched by age, sex, and health conditions. Models were stratified by sex, age, and deprivation.

**Results:** 23,428 TBIs (n=18,940) and 589,169 NTTs (n=421,259) were identified. TBIs were associated with higher mortality than NTT at all timepoints and age bands. Older age was associated with high mortality after TBI, with 16.9% of patients aged 65-79 (Hazard Ratio (HR)=4.52, 95% CI=[4.05,5.05], p<0.0001, relative to patients aged 18-39) and 31% of patients aged 80-100 dead within one month (9.08 [8.17,10.1], p<0.0001).

The impact of pre-injury dementia on post-TBI mortality was specifically during the chronic phase, 6 and 12 months after injury. Using NTT without dementia as a comparator, 12-month mortality was significantly higher in TBI patients with pre-injury dementia (2.15 [2.03,2.28], p<0.0001) than those without (1.52 [1.44,1.60], p<0.0001). Conversely, 30-day mortality was significant after TBI irrespective of dementia: TBI with (2.67 [2.42, 2.95], p<0.0001) or without pre-injury dementia (2.87 [2.65, 3.12], p<0.0001) had more than twice the mortality risk relative to NTT without dementia.

**Conclusions and relevance:** TBI is associated with higher all-cause mortality than NTT, particularly in older age. Pre-injury dementia was associated with particularly high mortality at 6 and 12 months. There is an urgent need to understand the reasons for poor outcomes in older adult TBI populations, especially those with pre-injury dementia.

## 1 Introduction

Traumatic brain injury (TBI) is a significant cause of death and disability worldwide^1,2^. The epidemiology of TBI has shifted towards older adults within the last two decades^3–5^. In high income countries, incidence of TBI and mortality from TBI in younger adults is showing relative decreases amid improved public health and road safety guidance^6–9^. In contrast, TBI incidence is increasing in older adults^6,10^. Cohort studies demonstrate that TBI in older adults is associated with higher short-term mortality compared to younger patients^10–15^, although mortality rates in older adults in the chronic setting after TBI (6+ months) has been less investigated.

Dementia is a common age-related comorbidity linked to high risk of falls^16,17^, an increasingly important cause of TBI^18,19^. Pre-injury dementia may also increase risk of poor outcomes post-TBI as neurodegenerative processes, such as neuroinflammation, may exacerbate secondary injury effects of TBI ^20,21^, while cognitive disturbances may interfere with rehabilitation and subsequent functional recovery. However, the impact of pre-existing dementia on TBI has not been systematically studied. Therefore, much is assumed, but little is known, about dementia and its effects on TBI outcomes.

Routinely collected electronic health record (EHR) data more broadly represent population-wide healthcare needs, enabling better observation of healthcare outcomes. First, the large sample minimises selection bias^22^, which allows more comprehensive and equitable assessment of TBI impact. It is possible to include higher representation from older, socio-economically deprived, or ethnic minority individuals, who may be disproportionately impacted by dementia^23,24^ and TBI^25^ but typically have low rates of participation and retention in prospective cohort studies^26–29^. Second, the longitudinal nature of EHRs facilitate longer follow-up, providing a much-needed view of post-TBI outcomes into the chronic phase. Third, the scale of population-based data enables detailed statistical matching, limiting potential confounding.

We examine all-cause mortality after hospitalised TBI, in older adults with and without existing dementia, within the Welsh population. Wales is one of four countries making up the United Kingdom with a population of three million^30^. Using primary and secondary care EHRs spanning 23 years, we compare the impacts of TBI compared to non-TBI trauma (NTT) across propensity-matched groups. We test the hypotheses that TBI is associated with higher mortality at one, six-, and twelve-months post-injury, an effect increased by older age. We then examine the combined impact of TBI and dementia, hypothesising that those with pre-injury dementia have higher mortality after TBI than those without.

## 2 Methods

### 2.1 Data

Linked EHRs were accessed through the Secure Anonymised Information Linkage (SAIL) Databank^31^ in Wales. Datasets included: Office of National Statistics (ONS) death data from the Annual District Death Extract (ADDE); hospital admissions from the Patient Episode Dataset for Wales (PEDW); demographic information from the Welsh Demographic Dataset (WDSD); GP consultations from the Welsh Longitudinal General Practice Dataset (WLGP), including data from 80% of practices in Wales.

### 2.2 Study Population

The study population included individuals with a hospital admission relating to TBI or NTT, aged 18-100 at injury, and registered with GP services in Wales from 1^st^ January 2000 to 31^st^ December 2022. Individuals entered the cohort on the date of TBI/NTT, and exited the study after the follow up periods of 1, 6, and 12 months or upon a date of death, whichever was sooner.

TBI and NTT were identified in hospital admissions using diagnostic codes from the international classification of diseases (ICD-10). TBI codes were validated for intracranial injury^32^. As no validated codes were available for NTT, patients were identified using codes for external injury (Chapter XIX: S00-T98)^33^. Among these, codes for intracranial and superficial injury were manually excluded. Head and skull injuries were included as TBI if intracranial injury was also recorded. All NTT/TBI codes were reviewed by a neurologist (LL) and are provided in Supplementary Tables 1-2.

To investigate the impact of dementia on post-injury outcomes, Individuals aged 40-100 were further categorised by their dementia status, creating four groups: dementia and TBI (DEM+TBI), dementia with NTT (DEM+NTT), non-dementia with TBI (NDEM+TBI), and non-dementia with NTT (NDEM+NTT). Dementia was identified in GP and hospital records using Read V2 and ICD-10 codes respectively (Supplementary Table 3).

Available demographic variables included week of birth, sex, and deprivation. Age was calculated by subtracting week of birth from the date of TBI/NTT and binned into groups (18-39, 40-64, 65-79, and 80-100). Deprivation was recorded as each individual’s Welsh Index of Multiple Deprivation (WIMD) quintile in 2019, with quintile one (Q1) being the most deprived. WIMD is a composite score describing deprivation across eight domains, including housing, education, and access to services^34^.

### 2.3 Event Clustering

TBI and NTT codes were clustered temporally to account for repeated coding within the same hospital admission. All diagnostic codes occurring within seven days were considered part of the same episode, while a diagnostic code occurring more than seven days after the index event was considered a new episode.

### 2.4 Mapping to mechanism

Injury mechanisms were identified by intersecting TBI/NTT codes with ICD-10 codes pertaining to assault, road traffic collision (RTC), or falls (Supplementary Table 4). Mechanism codes within seven days of the TBI/NTT were considered associated with the injury.

### 2.5 Statistical analysis

Python 3.11.4^35^ was used with the following packages: pyodbc, numpy, and pandas for data processing, scipy for statistical analysis, lifelines, statsmodels, and pingouin for Cox regression, tableone for summary statistics, and matplotlib for data visualisation.

### 2.6 Matching variables

Age, sex, and health conditions were used to match patients across groups. Health conditions included any previous diagnosis of common age-related conditions, including psychiatric (anxiety and depression), neurodegenerative non-dementia (Parkinson’s disease (PD), motor neuron disease (MND)), diabetes, cancer, respiratory (chronic obstructive pulmonary disease (COPD)), cardiovascular (hypertension, ischemic heart disease, congestive heart failure), liver (alcoholic/ non-alcoholic), renal (chronic kidney disease), and cerebrovascular conditions.

### 2.7 Propensity sampling and propensity matching

Logistic regression was used to calculate propensity scores using the specified matching variables. Age was standardised, and categorical variables (sex, morbidities) were converted into binary dummy variables. Propensity scores were computed as the predicted log odds (logit) from the logistic regression model for each individual.

The linear sum assignment algorithm was applied to find optimal matches between majority and minority group individuals. Groups were matched by minimising the sum of the absolute differences in their propensity scores, ensuring that matched individuals had similar propensity scores and distributions of the specified variables.

For the trauma and dementia analysis, groups were subset to individuals aged 40 and over. The three largest (majority) groups (DEM+NTT, NDEM+TBI, NDEM+NTT) were matched to the smallest (minority) group (DEM+TBI) using the same covariates in the following ratios: 2:1 NDEM+TBI, 5:1 DEM+NTT, 8:1 NDEM+NTT.

### 2.8 Survival analysis

Cox proportional hazards models were applied to estimate survival probability across the matched TBI/NTT groups, with the primary exposure being TBI/NTT injury.

Events were right-censored, i.e., with TBI/NTT as the known lower limit, and censorship occurring where subjects survived past the follow-up period (1, 6, or 12 months).

Models were adjusted for sex, age bands (18-39, 40-64, 65-79, 80-100), and WIMD quintile. Individuals with information missing in these variables were excluded.

We then modelled survival in the age, sex, and multimorbidity-matched groups aged 40-100 (see: section 4.6-4.7) with and without pre-injury dementia. Age bands, sex, and WIMD quintile were used as covariates.

## 3 Results

### 3.1 Cohort description

There were 4,001,332 individuals aged 18-100 and living in Wales from January 2000-December 2022 (Figure 1).

**Figure 1:**
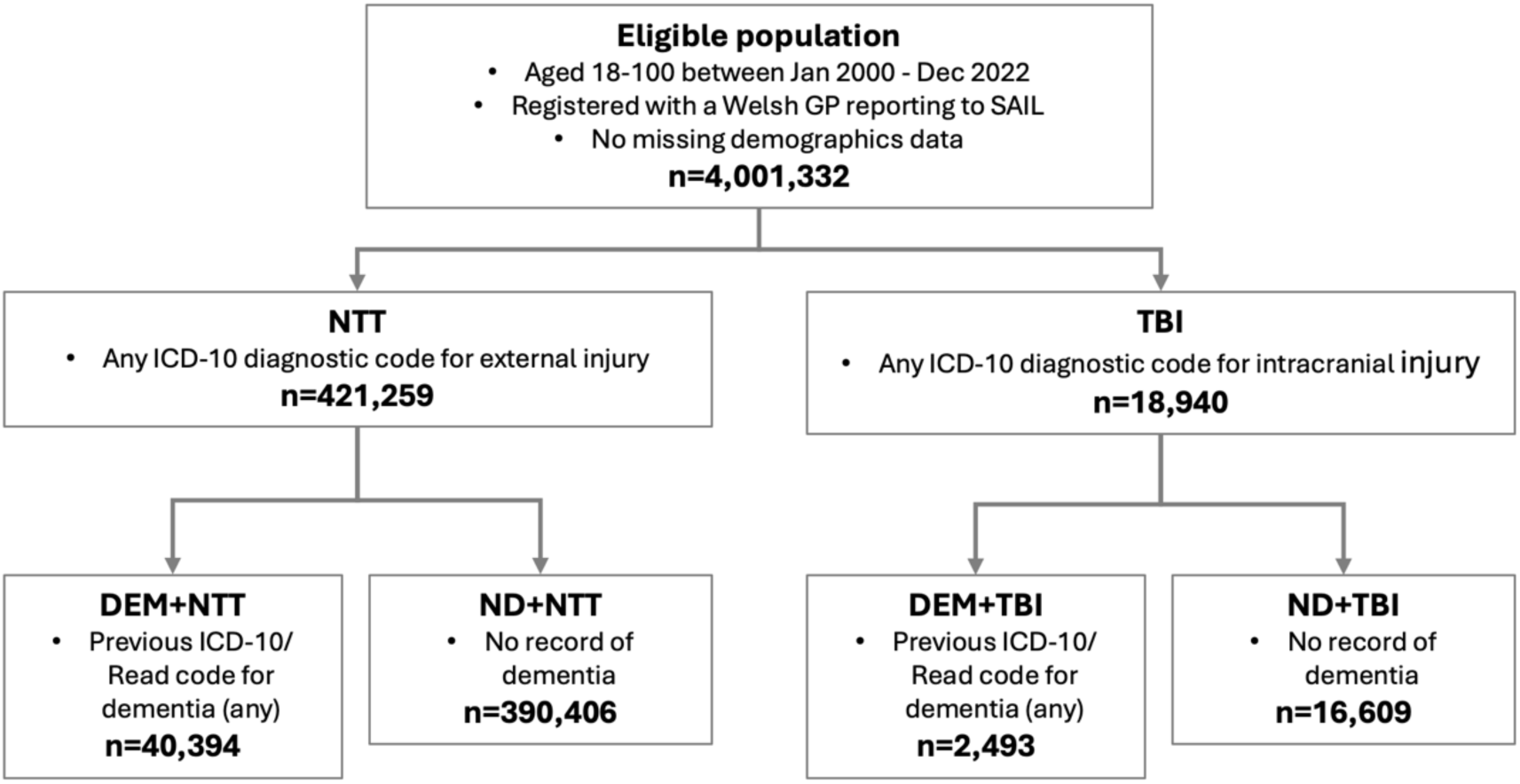
Flow diagram depicting total eligible population throughout the study period and individuals who were included in the subsequent analyses for the effects of injury (TBI vs NTT), followed by the effects of injury and pre-existing dementia (DEM vs ND). As the numbers for people with and without dementia are inclusive of individuals who sustained multiple injuries before and after their diagnosis of dementia, the events represented by each group (DEM+NTT, ND+NTT, DEM+TBI, ND+TBI) are unique, while some individuals are represented in multiple groups.

Over the 23-year study period, we identified 23,428 hospitalised TBI episodes across n=18,940 patients (Table 1) (Figure 2a-b), and 589,169 hospitalised NTT episodes in n=421,259. The most frequently recorded TBI diagnosis was traumatic subdural haemorrhage, and the most frequently recorded NTT diagnosis was neck of femur fracture. TBI/NTT diagnostic codes and counts are provided in Supplementary Tables 6-7.

**Figure 2:**
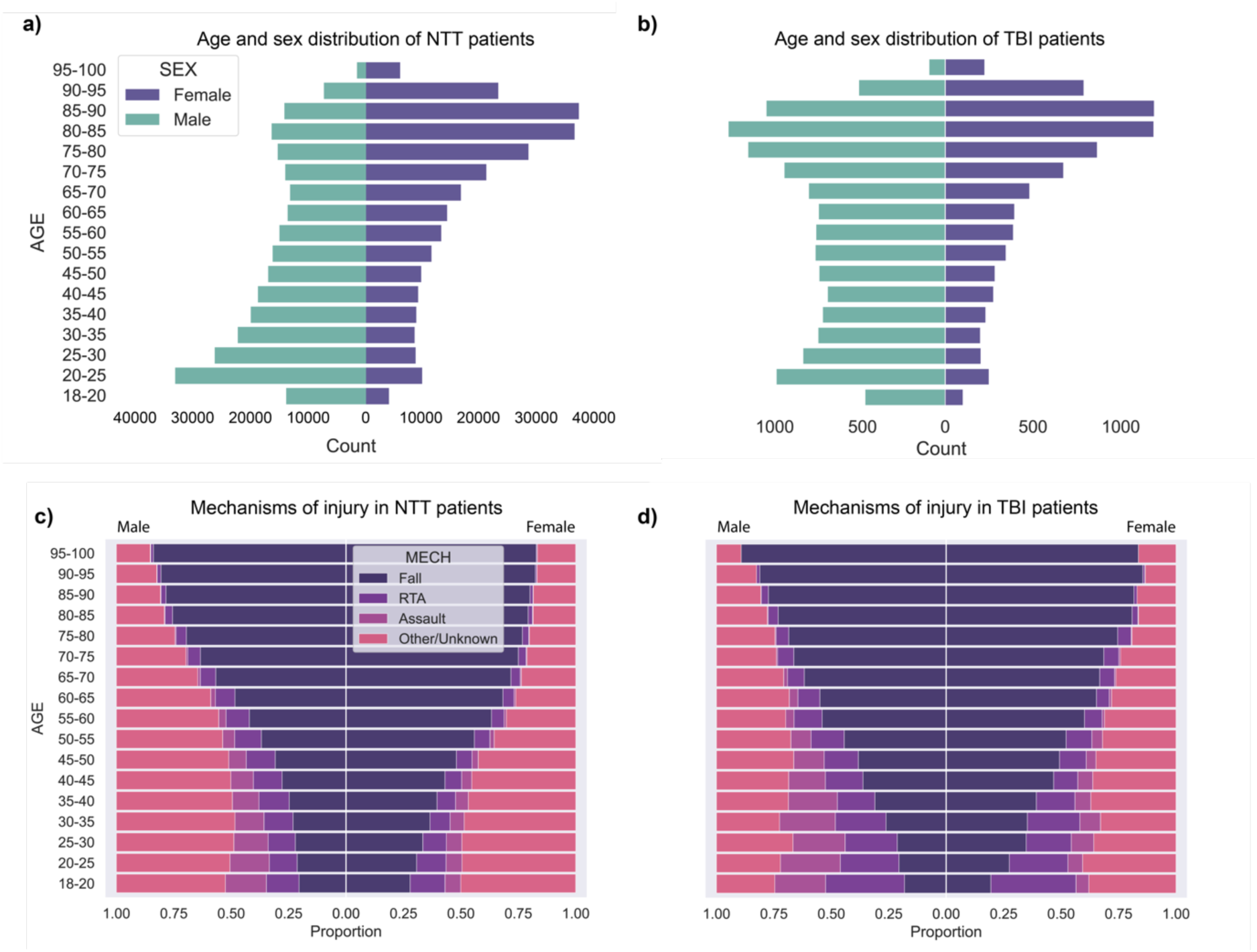
Demographics and injury mechanisms of NTT and TBI patients examined in this study. Males and females are mirrored across all plots, with males on the left and females on the right. **a-b)** Counts of **(a)** NTT and **(b)** TBI patients. Y-axis is shared across both plots and depicts five-year age bands, with the youngest group (18-20) at the bottom and the oldest group (95-100) at the top. X-axis depicts counts and differs across the NTT and TBI groups. **c-d)** Mechanisms associated with **c)** NTT and **d)** TBI, expressed in proportions per sex and age band. Both axes are shared across the plots: Y-axis again depicts five-year age bands, with the youngest at the bottom; X-axis depicts proportions of episodes associated with the following mechanisms: Falls (Indigo), road traffic collisions (RTCs; dark purple), assault (light purple), and all other or unknown causes (pink).

**Table 1:** Demographics of study population. For sample characteristics after matching, please see Supplementary Table 8.

|  |  | Overall | Non-TBI Trauma | TBI |
| --- | --- | --- | --- | --- |
| Episodes | - | 612,597 | 589,169 | 23,428 |
| N | - | 440,199 | 421,259 | 18,940 |
| Age, mean (SD) | - | 59.6 (23.8) | 59.5 (23.9) | 62.6 (22.7) |
| Sex, n (%) | Female | 304,618 (49.7) | 295,490 (50.2) | 9,128 (39.0) |
|  | Male | 307,978 (50.3) | 293,678 (49.8) | 14,300 (61.0) |
| Region, n (%) | Rural | 174,584 (30.1) | 167,947 (30.1) | 6,637 (30.0) |
|  | Urban | 404,718 (69.9) | 389,212 (69.9) | 15,506 (70.0) |
| Welsh Index of Multiple Deprivation (WIMD) Quintile (2019), n (%) | 1 (Most deprived) | 137,485 (22.4) | 132,263 (22.5) | 5,222 (22.3) |
|  | 2 | 132,278 (21.6) | 127,367 (21.6) | 4,911 (21.0) |
|  | 3 | 124,564 (20.3) | 119,931 (20.4) | 4,633 (19.8) |
|  | 4 | 114,468 (18.7) | 109,960 (18.7) | 4,508 (19.2) |
|  | 5 (Least deprived) | 103,752 (16.9) | 99,598 (16.9) | 4,154 (17.7) |
| Mechanism of injury, n (%) | Assault | 27,975 (4.6) | 26,595 (4.5) | 1,380 (5.9) |
|  | Fall | 306,270 (50.0) | 293,929 (49.9) | 12,341 (52.7) |
|  | Other/Unknown | 237,769 (38.8) | 230,291 (39.1) | 7,478 (31.9) |
|  | Road traffic collision (RTC) | 40,583 (6.6) | 38,354 (6.5) | 2,229 (9.5) |
| Discharge Destination, n (%) | Death | 25,607 (4.2) | 22,545 (3.8) | 3,062 (13.1) |
|  | Leaving medical care | 488,253 (79.8) | 473,799 (80.6) | 14,454 (61.8) |
|  | Ongoing medical care | 78,456 (12.8) | 73,416 (12.5) | 5,040 (21.5) |
|  | Other/Unknown | 729 (0.1) | 697 (0.1) | 32 (0.1) |
|  | Social/care home | 18,497 (3.0) | 17,686 (3.0) | 811 (3.5) |
| Comorbidities, n (%) | Psychiatric | 140,732 (23.0) | 134,525 (22.8) | 6,207 (26.5) |
|  | Neurodegenerative (non-dementia) | 12,199 (2.0) | 11,706 (2.0) | 493 (2.1) |
|  | Diabetes | 118,403 (19.3) | 112,832 (19.2) | 5,571 (23.8) |
|  | Cancer | 179,127 (29.2) | 171,504 (29.1) | 7,623 (32.5) |
|  | Respiratory | 68,599 (11.2) | 66,152 (11.2) | 2,447 (10.4) |
|  | Cardiovascular | 320,832 (52.4) | 307,397 (52.2) | 13,435 (57.3) |
|  | Liver | 18,741 (3.1) | 17,530 (3.0) | 1,211 (5.2) |
|  | Renal | 20,099 (3.3) | 19,130 (3.2) | 969 (4.1) |
|  | Cerebrovascular | 148,661 (24.3) | 140,697 (23.9) | 7,964 (34.0) |

Cohort characteristics are presented in Table 1, Supplementary Results 1, and Supplementary Table 8. There were more TBIs in younger males (18-39); however, TBI numbers were comparable between males and females in older age (Figure 2b). The majority of younger-age NTTs were in males, with the pattern reversing in older age (Figure 2a).

Characteristics of the cohorts with and without dementia are presented in Supplementary Tables 9-10 and Supplementary Results 2. The dementia groups had more fall-related injuries (DEM+NTT: 69.5%, DEM+TBI: 70.1%) than the non-dementia group (ND+NTT: 57.4%, ND+TBI: 58.2%), which had higher rates of RTC (ND+NTT: 5.2%, ND+TBI: 7.0%; DEM+NTT: 0.7%, DEM+TBI: 0.9%).

### 3.2 Falls, road traffic collisions and assaults are common causes of injury, but mechanism varies with age

Falls, road traffic collisions (RTCs), and assaults were common causes of TBI, but their relative frequencies were strongly dependent on age (Figure 2c-d). Among adults aged ≥55, falls were the most common mechanism of injury. This pattern was consistent across both sexes, with falls associated with 60% of TBIs in women and 54% in men aged 55-60. In those aged 85-100, this increased to 82% in women and 77% in men. Similar trends were observed for NTT (Supplementary Results 3, Supplementary Figures 1-2). After matching by age, sex, and multimorbidity, mechanisms were similar between TBI/NTT (Supplementary figures 3-4).

### 3.3 TBI, older age and greater deprivation associated with higher mortality across all timepoints

We next examined the impact of TBI. Across matched groups, TBI was associated with higher mortality risk relative to NTT at all timepoints (1M: 3.30 [3.17,3.44]; 6M: 2.00 [1.94,2.06]; 1Y: 1.76 [1.71,1.81]) (Figure 3a-c, hazard ratio plots).

**Figure 3:**
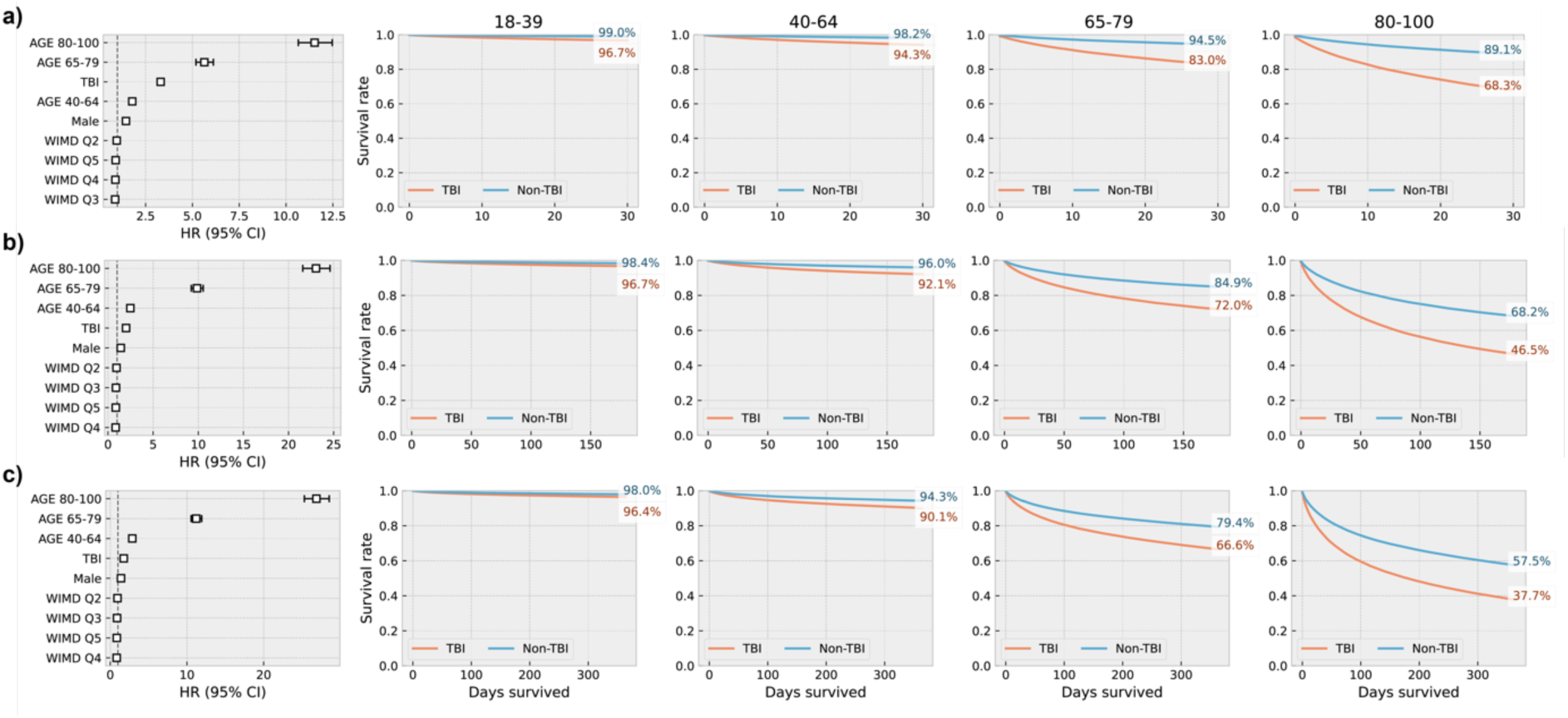
Survival probability at **a)** one month, **b)** six months, and **c)** twelve months across matched groups. Panels are organised as follows (left to right): Hazard ratios (HRs) for impact of examined factors on survival probability; survival curves across clinical groups in individuals aged 18-39; 40-64; 65-79; and 80-100. NB: Comparisons for HRs as follows: age bands (80-100, 65-79, and 40-64, compared to 18-39); clinical group (non-TBI trauma (NTT) compared to TBI); sex (males compared to females), and deprivation (Welsh Index for Multiple Deprivation (WIMD), with quintiles 2-5 compared to the most deprived quintile (Q1)).

Older age was associated with higher mortality for both TBI and NTT, with greater impact at chronic time points. The oldest group (80-100) was associated with the highest mortality risk. At one month, mortality risk in this group was 11x that of the youngest group (18-39) (11.5, [10.7,12.5]) (Figure 3a), increasing over time (6M: 23.0, [21.6,24.6]; 12M: 26.9, [25.3,28.5]) (Figures 3b-c).

The 65-79 age band was associated with 5-11x the risk of mortality at one month (1M: 5.63 [5.19,6.12]; 6M: 9.86 [9.22,10.6]; 12M: 11.2 [10.5,11.9]).

The 40-64 age band was also associated with significantly higher mortality, between 1-3x times that of the youngest group (1M: 1.78 [1.63,1.96]; 6M: 2.47 [2.30,2.67]; 12M: 2.86 [2.68,3.07]).

Male sex and greater deprivation were generally associated with higher mortality. All p-values were significant at p<0.0001. Full statistical results are in Supplementary Tables 11-16, Supplementary Results 4.

### 3.4 All-cause mortality is highest in TBI with pre-injury dementia at chronic timepoints

We next quantified the impact of pre-injury dementia on mortality after TBI. At one month, the TBI groups had the highest mortality irrespective of pre-injury dementia. The NDEM+TBI group had the highest mortality (2.87 [2.65,3.12]), with similarly high mortality in the DEM+TBI group (2.67 [2.42,2.95]) (Figure 4a hazard ratio plot).

**Figure 4:**
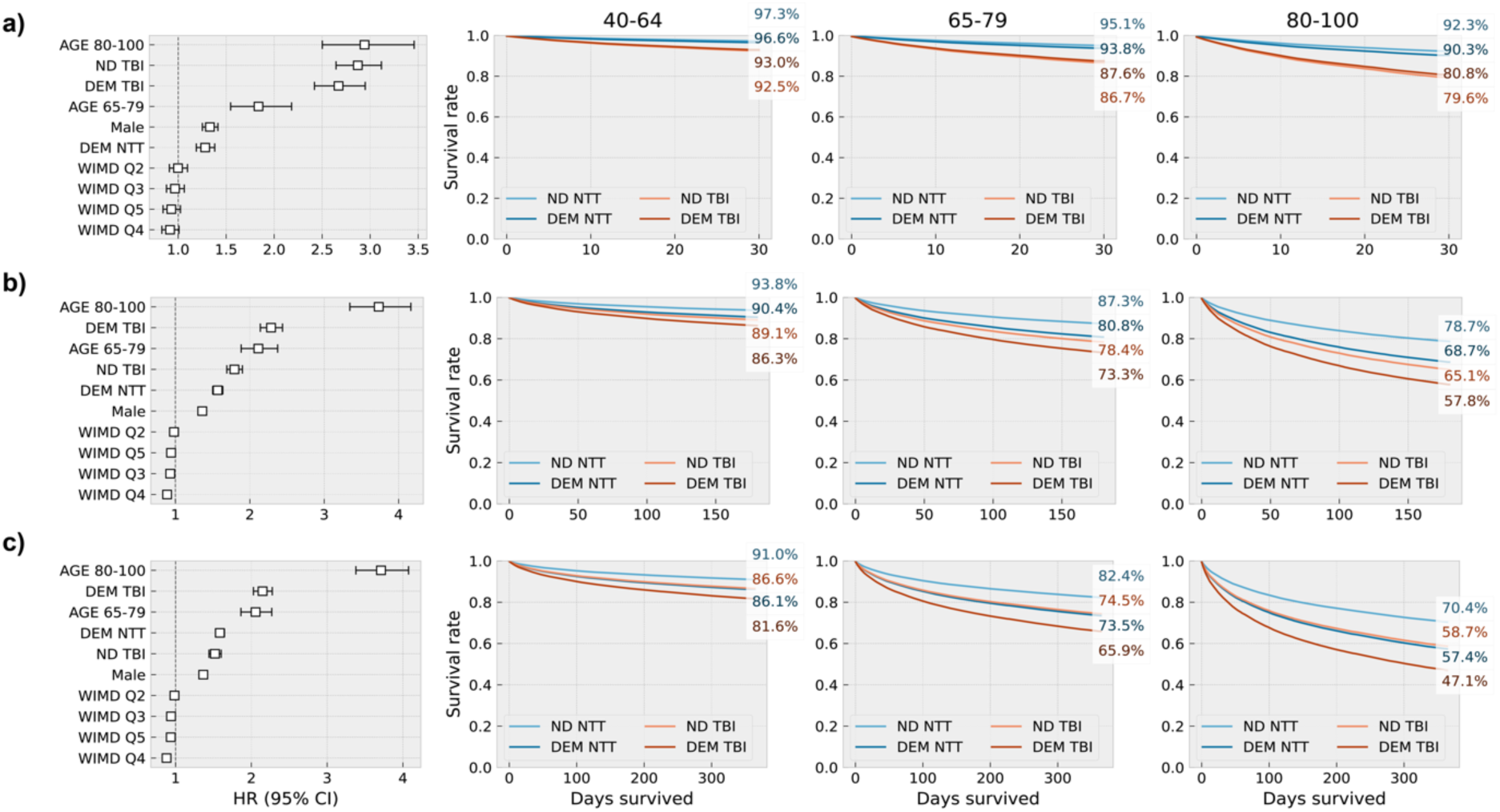
Cox regression results and survival curves at **a)** one month, **b)** six months, and **c)** twelve months post-injury across matched samples. Panels are organised as follows (left to right): Hazard ratios (HRs) for impact of examined factors on survival probability, survival curves across clinical groups in individuals aged 40-64; 65-79; and 80-100. NB: Comparisons for HRs as follows: age bands (80-100 and 65-79, compared to 40-64); clinical group (Dementia and TBI (DEM-TBI), dementia and NTT (DEM-NTT), non-dementia and TBI (ND-TBI), all compared to non-dementia and NTT (ND-NTT); sex (males compared to females), and deprivation quintiles (Welsh Index for Multiple Deprivation (WIMD), quintiles 1-5, with quintiles 2-5 compared to the most deprived quintile (Q1)).

By six months and one year post-injury, mortality was highest in the DEM+TBI group (6M: 2.29 [2.14,2.44]; 1Y: 2.15 [2.03,2.28]), followed by NDEM+TBI (6M: 1.79 [1.69,1.90]; 1Y: 1.52 [1.44,1.60]) (Figures 4b-c). The DEM+NTT group was also associated with higher mortality than the NDEM+NTT group at all timepoints (DEM+NTT: 1M: 1.28 [1.19,1.38]; 6M: 1.57 [1.50,1.64]; 12M: 1.59 [1.53,1.65]).

Age effects remained significant within this matched sample, with the oldest age group (80-100) having a threefold risk of mortality relative to those aged 40-64 at 1 (2.94 [2.50,3.46]), 6 (3.73 [3.35,4.17]), and 12 months (3.71 [3.38,4.07]). Individuals aged 65-79 had approximately double the mortality risk at 1 (1.84 [1.55,2.18]), 6 (2.11 [1.88,2.37]), and 12 months (2.06 [1.86,2.27]).

Male sex was generally associated with higher mortality, and greater deprivation was associated with higher mortality from 6-12 months. For full statistical results, please see Supplementary Tables 17-22, Supplementary Results 5.

## 4 Discussion

We systematically study the impacts of older age, and pre-injury dementia, a common comorbidity in older adults, on mortality after hospitalised TBI. We confirm, replicating at scale, two findings from the literature: TBI is associated with greater mortality than NTT^36^, and older adults have much higher mortality after TBI^10–15^. Our novel finding, in the largest population study to date with careful matching for comorbidities, is that existing dementia disproportionately impacts post-TBI mortality. This effect exacerbates with age and is particularly marked at chronic timepoints.

Older adults with dementia may have worse outcomes after TBI for several reasons. We found that the increased mortality in comorbid TBI and dementia is only apparent from 6 months post-TBI, suggesting that the impact of dementia on post-TBI mortality may not be through acute factors such as increased injury extent. TBI is a risk factor for neurodegeneration^37^, and there is an established link between TBI and later onset of dementia, associated with a shared mechanism of chronic microglial activation and subsequent neuroinflammation that potentially accelerates existing neurodegenerative pathology^38,39^. A TBI may set those with pre-injury dementia on a trajectory of steeper functional decline, the impact of which becomes increasingly evident with time. Another reason is that this population lives with significant multimorbidity^40^. While health conditions were used as matching factors in this study, people living with dementia may experience additional challenges in coping with the same conditions^41^, such as adhering to medication routines^42^ or engaging with rehabilitation^43^. Dementia may also delay the recognition and identification of symptoms during acute illness^44^, which may lead to inadequate management of post-TBI problems and modify re-injury risk.

Poor outcomes have been consistently observed with older age^45–48^. However, previous studies of hospitalised TBI have focused on acute mortality. Our findings are well-aligned with existing cohort studies reporting 16-30% mortality within 30 days in ages 65-79^5,12,46,49^ and 30% in ages 80-100^5,12^. We extend the literature by examining chronic survival post-injury, finding that 6- and 12-month mortality was significantly higher in patients aged 80-100 than patients aged 65-79. It is important to consider the extent to which older age and presence of dementia contributes to high post-TBI mortality because of inequalities in healthcare provision. Older patients are likely to experience longer delays between admission and imaging assessment, and less likely to be transferred for neurosurgical intervention^50^. Older adults, especially those with cognitive impairment, are also less likely to receive specialised rehabilitation services^51–56^, impeding functional recovery. Further, frail older patients may require specialised accommodations that may not be available in less-resourced settings^57^. These differences in care may lead to a self-fulling prophecy in the care of older TBI patients, where the provision of care and potential recovery are limited within this demographic as they are presumed to do poorly.

We also observed a significant association between old age and TBI occurrence, and that the male preponderance in TBI seen in young adults is not observed in older groups. TBIs in women are consistently understudied within the literature, which likely relates to the paucity of research in older adult TBI. Our results advocate for adequate older adult and female representation in future TBI research. We further showed that the vast majority of TBIs in those aged ≥85 across both sexes were due to falls. Falls, especially low-energy slips and trips at home, are driving increased older-age TBI in high income countries^58,59^. This has implications for clinical care, as presentation with a ‘mild’ TBI (often defined as a score of 14-15 on the Glasgow Coma Scale (GCS)^60^) from a low-energy mechanism in an older adult should still be treated with high suspicion for significant head injury^61^. This is now recognised within imaging guidelines^62,63^, with older age included as a risk factor for significant brain injury independently of mechanism or clinical presentation. Blood-based biomarkers that reflect injury extent, for example glial fibrillary acidic protein (GFAP)^64^, may help identify patients likely to have significant injury, although further work is needed to establish whether normative ranges derived from the current literature are valid in older adults^65^.

This finding also has implications for TBI prevention. Measures resulting in reduced TBIs in younger patients, e.g., encouraging seatbelt or helmet usage to limit RTC-related TBIs, are unlikely to substantially reduce the burden of older adult TBI. Amid improvements in care and life expectancy, older adults are living with more health conditions and, concurrently, a greater number of medications. Polypharmacy and multimorbidity, both linked to frailty^66–68^, are predictive of fall risk^69–71^, with medications including antidepressants and sedatives^70,72–74^ specifically identified as contributors. Long-term anticoagulant use is associated with post-traumatic intracranial haemorrhage^75^ and may contribute to TBI-related hospitalisation and subsequent mortality^76^. Future work should investigate how specific medications contribute to TBI risk and explore medication rationalisation as a measure to reduce TBIs and their consequences. Future work should also support falls prevention strategies such as exercise programmes^77^ and comprehensive geriatric assessment (CGA), which evaluates multiple domains (such as medication, cognition, and mobility)^78,79^.

We note several limitations to our study. Like others using routine electronic health records, our findings depend on the quality of coding, population characteristics, and geographical coverage of the EHRs. To assess the replicability and generalisability of our findings, our study should be reproduced using other EHR databases with different demographic and socio-economic profiles and systems of healthcare provision. Next, we identified TBI using validated diagnostic coding^32^, however we did not have a classification of severity. Likewise, we could not stratify dementia by disease stage, which may have independently impacted post-injury survival. Given the high proportion of polytrauma within this population, we were unable to evaluate the isolated impact of TBI. However, the inclusion of an NTT group mitigates potential bias due to trauma burden. Our population-based approach allowed us to perform detailed, individual-level matching for key confounds such as age, sex, and health conditions, giving us confidence that our findings reflect the specific impact of dementia. For example, while falls in people with dementia may be precipitated by clinical conditions (e.g. cardiovascular conditions) that can independently lead to high mortality, our matching enabled comparison between groups with similar profiles of multimorbidity at the point of injury.

In this population-based study of older adults in Wales, we observed substantial all-cause mortality following hospitalised TBI within the last twenty years, particularly in people with pre-injury dementia. Further research is needed to understand contributors to this disproportionate mortality, such as biological vulnerabilities or differences in care pathways for older and comorbid patients with TBI.

## Data Availability

Data from this study are held in the Secure Anonymised Information Linkage (SAIL) Databank at Swansea University; while we cannot share data directly, SAIL welcomes applications (https://saildatabank.com/application-process) to access this dataset for approved research purposes.

## Acknowledgements

We would like to express our gratitude to our colleagues in the UK Dementia Research Institute Care Research and Technology Centre (UK DRI CR&T) for their ongoing support and insightful feedback. We are also grateful to the team at the Secure Anonymised Information Linkage (SAIL) Databank for their assistance throughout this project.

## References

1. Maas AIR, Menon DK, Adelson PD, et al. Traumatic brain injury: integrated approaches to improve prevention, clinical care, and research. Lancet Neurol. 2017;16(12):987–1048. doi:10.1016/S1474-4422(17)30371-X

2. Maas AIR, Menon DK, Manley GT, et al. Traumatic brain injury: progress and challenges in prevention, clinical care, and research. Lancet Neurol. 2022;21(11):1004–1060. doi:10.1016/S1474-4422(22)00309-X

3. Dixon JR, Lecky F, Bouamra O, et al. Age and the distribution of major injury across a national trauma system. Age Ageing. 2020;49(2):218–226. doi:10.1093/ageing/afz151

4. Cusimano MD, Saarela O, Hart K, Zhang S, McFaull SR. A population-based study of fall-related traumatic brain injury identified in older adults in hospital emergency departments. Neurosurg Focus. 2020;49(4):E20. doi:10.3171/2020.7.FOCUS20520

5. Hawley C, Sakr M, Scapinello S, Salvo J, Wrenn P. Traumatic brain injuries in older adults—6 years of data for one UK trauma centre: retrospective analysis of prospectively collected data. Emerg Med J. 2017;34(8):509–516. doi:10.1136/emermed-2016-206506

6. Roozenbeek B, Maas AIR, Menon DK. Changing patterns in the epidemiology of traumatic brain injury. Nat Rev Neurol. 2013;9(4):231–236. doi:10.1038/nrneurol.2013.22

7. Maas AI, Stocchetti N, Bullock R. Moderate and severe traumatic brain injury in adults. Lancet Neurol. 2008;7(8):728–741. doi:10.1016/S1474-4422(08)70164-9

8. Taylor CA. Traumatic Brain Injury–Related Emergency Department Visits, Hospitalizations, and Deaths — United States, 2007 and 2013. MMWR Surveill Summ. 2017;66. doi:10.15585/mmwr.ss6609a1

9. Saxena S, Zutrauen S, McFaull SR. Assault-related traumatic brain injury hospitalizations in Canada from 2010 to 2021: rates, trends and comorbidity. Inj Epidemiol. 2024;11(1):4. doi:10.1186/s40621-024-00486-5

10. Marincowitz C, Lecky F, Allgar V, Sheldon T. Evaluation of the impact of the NICE head injury guidelines on inpatient mortality from traumatic brain injury: an interrupted time series analysis. BMJ Open. 2019;9(6):e028912. doi:10.1136/bmjopen-2019-028912

11. LeBlanc J, Guise E de, Gosselin N, Feyz M. Comparison of functional outcome following acute care in young, middle-aged and elderly patients with traumatic brain injury. Brain Inj. 2006;20(8):779–790. doi:10.1080/02699050600831835

12. Utomo WK, Gabbe BJ, Simpson PM, Cameron PA. Predictors of in-hospital mortality and 6-month functional outcomes in older adults after moderate to severe traumatic brain injury. Injury. 2009;40(9):973–977. doi:10.1016/j.injury.2009.05.034

13. Maiden MJ, Cameron PA, Rosenfeld JV, Cooper DJ, McLellan S, Gabbe BJ. Long-Term Outcomes after Severe Traumatic Brain Injury in Older Adults. A Registry-based Cohort Study. Am J Respir Crit Care Med. 2020;201(2):167–177. doi:10.1164/rccm.201903-0673OC

14. Hunt C, Zahid S, Ennis N, et al. Quality of life measures in older adults after traumatic brain injury: a systematic review. Qual Life Res. 2019;28(12):3137–3151. doi:10.1007/s11136-019-02297-4

15. Downing MG, Carty M, Olver J, et al. The impact of age on outcome 2 years after traumatic brain injury: Case control study. Ann Phys Rehabil Med. 2024;67(5):101834. doi:10.1016/j.rehab.2024.101834

16. Allan LM, Ballard CG, Rowan EN, Kenny RA. Incidence and Prediction of Falls in Dementia: A Prospective Study in Older People. PLOS ONE. 2009;4(5):e5521. doi:10.1371/journal.pone.0005521

17. Shaw FE. Falls in cognitive impairment and dementia. Clin Geriatr Med. 2002;18(2):159–173. doi:10.1016/S0749-0690(02)00003-4

18. Peeters W, van den Brande R, Polinder S, et al. Epidemiology of traumatic brain injury in Europe. Acta Neurochir (Wien). 2015;157(10):1683–1696. doi:10.1007/s00701-015-2512-7

19. Brazinova A, Rehorcikova V, Taylor MS, et al. Epidemiology of Traumatic Brain Injury in Europe: A Living Systematic Review. J Neurotrauma. 2021;38(10):1411–1440. doi:10.1089/neu.2015.4126

20. Collins-Praino LE, Corrigan F. Does neuroinflammation drive the relationship between tau hyperphosphorylation and dementia development following traumatic brain injury? Brain Behav Immun. 2017;60:369–382. doi:10.1016/j.bbi.2016.09.027

21. Pasqualetti G, Brooks DJ, Edison P. The Role of Neuroinflammation in Dementias. Curr Neurol Neurosci Rep. 2015;15(4):17. doi:10.1007/s11910-015-0531-7

22. Kho ME, Duffett M, Willison DJ, Cook DJ, Brouwers MC. Written informed consent and selection bias in observational studies using medical records: systematic review. BMJ. 2009;338:b866. doi:10.1136/bmj.b866

23. Kornblith E, Bahorik A, Boscardin WJ, Xia F, Barnes DE, Yaffe K. Association of Race and Ethnicity With Incidence of Dementia Among Older Adults. JAMA. 2022;327(15):1488–1495. doi:10.1001/jama.2022.3550

24. Pham TM, Petersen I, Walters K, et al. Trends in dementia diagnosis rates in UK ethnic groups: analysis of UK primary care data. Clin Epidemiol. 2018;10:949–960. doi:10.2147/CLEP.S152647

25. Maldonado J, Huang JH, Childs EW, Tharakan B. Racial/Ethnic Differences in Traumatic Brain Injury: Pathophysiology, Outcomes, and Future Directions. J Neurotrauma. 2023;40(5-6):502–513. doi:10.1089/neu.2021.0455

26. Gilmore-Bykovskyi AL, Jin Y, Gleason C, et al. Recruitment and retention of underrepresented populations in Alzheimer’s disease research: A systematic review. Alzheimers Dement Transl Res Clin Interv. 2019;5:751–770. doi:10.1016/j.trci.2019.09.018

27. Corrigan JD, Harrison-Felix C, Bogner J, Dijkers M, Terrill MS, Whiteneck G. Systematic bias in traumatic brain injury outcome studies because of loss to follow-up. Arch Phys Med Rehabil. 2003;84(2):153–160. doi:10.1053/apmr.2003.50093

28. Sander AM, Lequerica AH, Ketchum JM, et al. Race/Ethnicity and Retention in Traumatic Brain Injury Outcomes Research: A Traumatic Brain Injury Model Systems National Database Study. J Head Trauma Rehabil. 2018;33(4):219–227. doi:10.1097/HTR.0000000000000395

29. Vyas MV, Raval PK, Watt JA, Tang-Wai DF. Representation of ethnic groups in dementia trials: systematic review and meta-analysis. J Neurol Sci. 2018;394:107–111. doi:10.1016/j.jns.2018.09.012

30. Population and household estimates, Wales - Office for National Statistics. Accessed April 24, 2024. https://www.ons.gov.uk/peoplepopulationandcommunity/populationandmigration/populationestimates/bulletins/populationandhouseholdestimateswales/census2021

31. Lyons RA, Jones KH, John G, et al. The SAIL databank: linking multiple health and social care datasets. BMC Med Inform Decis Mak. 2009;9(1):3. doi:10.1186/1472-6947-9-3

32. Gabella BA, Hathaway JE, Hume B, et al. Multisite medical record review of emergency department visits for traumatic brain injury. Inj Prev. 2021;27(Suppl 1):i42–i48. doi:10.1136/injuryprev-2019-043510

33. World Health Organisation. International Statistical Classification of Diseases and Related Health Problems 10th Revision - ICD-10 Version: 2019. Accessed November 17, 2022. https://icd.who.int/browse10/2019/en

34. Gov.wales. Welsh Index of Multiple Deprivation (full Index update with ranks): 2019. GOV.WALES. Accessed February 9, 2022. https://gov.wales/welsh-index-multiple-deprivation-full-index-update-ranks-2019

35. Python Release Python 3.11.4. Python.org. Accessed June 12, 2024. https://www.python.org/downloads/release/python-3114/

36. Elser H, Gottesman RF, Walter AE, et al. Head Injury and Long-term Mortality Risk in Community-Dwelling Adults. JAMA Neurol. 2023;80(3):260–269. doi:10.1001/jamaneurol.2022.5024

37. Brett BL, Gardner RC, Godbout J, Dams-O’Connor K, Keene CD. Traumatic Brain Injury and Risk of Neurodegenerative Disorder. Biol Psychiatry. 2022;91(5):498–507. doi:10.1016/j.biopsych.2021.05.025

38. Faden AI, Loane DJ. Chronic Neurodegeneration After Traumatic Brain Injury: Alzheimer Disease, Chronic Traumatic Encephalopathy, or Persistent Neuroinflammation? Neurotherapeutics. 2015;12(1):143–150. doi:10.1007/s13311-014-0319-5

39. Gardner RC, Burke JF, Nettiksimmons J, Kaup A, Barnes DE, Yaffe K. Dementia Risk After Traumatic Brain Injury vs Nonbrain Trauma: The Role of Age and Severity. JAMA Neurol. 2014;71(12):1490–1497. doi:10.1001/jamaneurol.2014.2668

40. Schubert CC, Boustani M, Callahan CM, et al. Comorbidity Profile of Dementia Patients in Primary Care: Are They Sicker? J Am Geriatr Soc. 2006;54(1):104–109. doi:10.1111/j.1532-5415.2005.00543.x

41. Bunn F, Burn AM, Robinson L, et al. Healthcare organisation and delivery for people with dementia and comorbidity: a qualitative study exploring the views of patients, carers and professionals. BMJ Open. 2017;7(1):e013067. doi:10.1136/bmjopen-2016-013067

42. Lim RH, Sharmeen T. Medicines management issues in dementia and coping strategies used by people living with dementia and family carers: A systematic review. Int J Geriatr Psychiatry. 2018;33(12):1562–1581. doi:10.1002/gps.4985

43. van der Wardt V, Hancox J, Gondek D, et al. Adherence support strategies for exercise interventions in people with mild cognitive impairment and dementia: A systematic review. Prev Med Rep. 2017;7:38–45. doi:10.1016/j.pmedr.2017.05.007

44. Fox C, Smith T, Maidment I, et al. The importance of detecting and managing comorbidities in people with dementia? Age Ageing. 2014;43(6):741–743. doi:10.1093/ageing/afu101

45. Testa JA, Malec JF, Moessner AM, Brown AW. Outcome After Traumatic Brain Injury: Effects of Aging on Recovery. Arch Phys Med Rehabil. 2005;86(9):1815–1823. doi:10.1016/j.apmr.2005.03.010

46. Mosenthal AC, Lavery RF, Addis M, et al. Isolated Traumatic Brain Injury: Age Is an Independent Predictor of Mortality and Early Outcome. J Trauma Acute Care Surg. 2002;52(5):907–911.

47. Flaada JT, Leibson CL, Mandrekar JN, et al. Relative Risk of Mortality after Traumatic Brain Injury: A Population-Based Study of The Role of Age And Injury Severity. J Neurotrauma. 2007;24(3):435–445. doi:10.1089/neu.2006.0119

48. Stocchetti N, Paternò R, Citerio G, Beretta L, Colombo A. Traumatic Brain Injury in an Aging Population. J Neurotrauma. 2012;29(6):1119–1125. doi:10.1089/neu.2011.1995

49. Rozzelle CJ, Wofford JL, Branch CL. Predictors of hospital mortality in older patients with subdural hematoma. J Am Geriatr Soc. 1995;43(3):240–244. doi:10.1111/j.1532-5415.1995.tb07329.x

50. Kirkman MA, Jenks T, Bouamra O, Edwards A, Yates D, Wilson MH. Increased mortality associated with cerebral contusions following trauma in the elderly: bad patients or bad management? J Neurotrauma. 2013;30(16):1385–1390. doi:10.1089/neu.2013.2881

51. Waltzman D, Haarbauer-Krupa J, Womack LS. Traumatic Brain Injury in Older Adults—A Public Health Perspective. JAMA Neurol. 2022;79(5):437–438. doi:10.1001/jamaneurol.2022.0114

52. Gardner RC, Dams-O’Connor K, Morrissey MR, Manley GT. Geriatric Traumatic Brain Injury: Epidemiology, Outcomes, Knowledge Gaps, and Future Directions. J Neurotrauma. 2018;35(7):889–906. doi:10.1089/neu.2017.5371

53. Klang A, Molero Y, Lichtenstein P, et al. Access to Rehabilitation After Hospitalization for Traumatic Brain Injury: A National Longitudinal Cohort Study in Sweden. Neurorehabil Neural Repair. 2023;37(11-12):763–774. doi:10.1177/15459683231209315

54. Thompson HJ, Weir S, Rivara FP, et al. Utilization and Costs of Health Care after Geriatric Traumatic Brain Injury. J Neurotrauma. 2012;29(10):1864–1871. doi:10.1089/neu.2011.2284

55. McGilton KS, Davis AM, Naglie G, et al. Evaluation of patient-centered rehabilitation model targeting older persons with a hip fracture, including those with cognitive impairment. BMC Geriatr. 2013;13(1):136. doi:10.1186/1471-2318-13-136

56. Isbel ST, Jamieson MI. Views from health professionals on accessing rehabilitation for people with dementia following a hip fracture. Dementia. 2017;16(8):1020–1031. doi:10.1177/1471301216631141

57. Galimberti S, Graziano F, Maas AIR, et al. Effect of frailty on 6-month outcome after traumatic brain injury: a multicentre cohort study with external validation. Lancet Neurol. 2022;21(2):153–162. doi:10.1016/S1474-4422(21)00374-4

58. Kehoe A, Smith JE, Edwards A, Yates D, Lecky F. The changing face of major trauma in the UK. Emerg Med J EMJ. 2015;32(12):911–915. doi:10.1136/emermed-2015-205265

59. Rickard F, Gale J, Williams A, Shipway D. New horizons in subdural haematoma. Age Ageing. 2023;52(12):afad240. doi:10.1093/ageing/afad240

60. Malec JF, Brown AW, Leibson CL, et al. The Mayo Classification System for Traumatic Brain Injury Severity. J Neurotrauma. 2007;24(9):1417–1424. doi:10.1089/neu.2006.0245

61. Lecky FE, Otesile O, Marincowitz C, et al. The burden of traumatic brain injury from low-energy falls among patients from 18 countries in the CENTER-TBI Registry: A comparative cohort study. PLOS Med. 2021;18(9):e1003761. doi:10.1371/journal.pmed.1003761

62. Stiell IG, Wells GA, Vandemheen K, et al. The Canadian CT Head Rule for patients with minor head injury. Lancet Lond Engl. 2001;357(9266):1391-1396. doi:10.1016/s0140-6736(00)04561-x

63. Overview | Head injury: assessment and early management | Guidance | NICE. May 18, 2023. Accessed May 2, 2024. https://www.nice.org.uk/guidance/ng232

64. Abdelhak A, Foschi M, Abu-Rumeileh S, et al. Blood GFAP as an emerging biomarker in brain and spinal cord disorders. Nat Rev Neurol. 2022;18(3):158–172. doi:10.1038/s41582-021-00616-3

65. Gardner RC, Puccio AM, Korley FK, et al. Effects of age and time since injury on traumatic brain injury blood biomarkers: a TRACK-TBI study. Brain Commun. 2023;5(1):fcac316. doi:10.1093/braincomms/fcac316

66. Hoogendijk EO, Afilalo J, Ensrud KE, Kowal P, Onder G, Fried LP. Frailty: implications for clinical practice and public health. The Lancet. 2019;394(10206):1365–1375. doi:10.1016/S0140-6736(19)31786-6

67. Vetrano DL, Palmer K, Marengoni A, et al. Frailty and Multimorbidity: A Systematic Review and Meta-analysis. J Gerontol Ser A. 2019;74(5):659-666. doi:10.1093/gerona/gly110

68. Veronese N, Stubbs B, Noale M, et al. Polypharmacy Is Associated With Higher Frailty Risk in Older People: An 8-Year Longitudinal Cohort Study. J Am Med Dir Assoc. 2017;18(7):624–628. doi:10.1016/j.jamda.2017.02.009

69. Rubenstein LZ. Falls in older people: epidemiology, risk factors and strategies for prevention. Age Ageing. 2006;35(suppl_2):ii37-ii41. doi:10.1093/ageing/afl084

70. Richardson K, Bennett K, Kenny RA. Polypharmacy including falls risk-increasing medications and subsequent falls in community-dwelling middle-aged and older adults. Age Ageing. 2015;44(1):90–96. doi:10.1093/ageing/afu141

71. Dhalwani NN, Fahami R, Sathanapally H, Seidu S, Davies MJ, Khunti K. Association between polypharmacy and falls in older adults: a longitudinal study from England. BMJ Open. 2017;7(10):e016358. doi:10.1136/bmjopen-2017-016358

72. Thorell K, Ranstad K, Midlöv P, Borgquist L, Halling A. Is use of fall risk-increasing drugs in an elderly population associated with an increased risk of hip fracture, after adjustment for multimorbidity level: a cohort study. BMC Geriatr. 2014;14(1):131. doi:10.1186/1471-2318-14-131

73. Kerse N, Flicker L, Pfaff JJ, et al. Falls, Depression and Antidepressants in Later Life: A Large Primary Care Appraisal. PLOS ONE. 2008;3(6):e2423. doi:10.1371/journal.pone.0002423

74. Marcum ZA, Perera S, Thorpe JM, et al. Antidepressant Use and Recurrent Falls in Community-Dwelling Older Adults: Findings From the Health ABC Study. Ann Pharmacother. 2016;50(7):525–533. doi:10.1177/1060028016644466

75. Pieracci FM, Eachempati SR, Shou J, Hydo LJ, Barie PS. Use of Long-Term Anticoagulation is Associated With Traumatic Intracranial Hemorrhage and Subsequent Mortality in Elderly Patients Hospitalized After Falls: Analysis of the New York State Administrative Database. J Trauma Acute Care Surg. 2007;63(3):519. doi:10.1097/TA.0b013e31812e519b

76. Inui TS, Parina R, Chang DC, Inui TS, Coimbra R. Mortality after ground-level fall in the elderly patient taking oral anticoagulation for atrial fibrillation/flutter: A long-term analysis of risk versus benefit. J Trauma Acute Care Surg. 2014;76(3):642. doi:10.1097/TA.0000000000000138

77. Sherrington C, Fairhall NJ, Wallbank GK, et al. Exercise for preventing falls in older people living in the community. Cochrane Database Syst Rev. 2019;(1). doi:10.1002/14651858.CD012424.pub2

78. Veronese N, Custodero C, Demurtas J, et al. Comprehensive geriatric assessment in older people: an umbrella review of health outcomes. Age Ageing. 2022;51(5):afac104. doi:10.1093/ageing/afac104

79. Hartley P, Forsyth F, Rowbotham S, Briggs R, Kenny RA, Romero-Ortuno R. The use of the World Guidelines for Falls Prevention and Management’s risk stratification algorithm in predicting falls in The Irish Longitudinal Study on Ageing (TILDA). Age Ageing. 2023;52(7):afad129. doi:10.1093/ageing/afad129

