## Supplementary Materials for "Traumatic brain injury (TBI) and mortality in older adults with and without pre-injury dementia"


| Index | Title | Page(s) |
|---|---|---|
| **Tables** | | |
| 1 | ICD-10 codes for traumatic brain injury (TBI) | 2 |
| 2 | ICD-10 codes for non-TBI trauma (NTT) | 3-4 |
| 3 | ICD-10 and Read V2 codes for dementia | 5-6 |
| 4 | ICD-10 codes for injury mechanisms | 7-10 |
| 5 | Mappings for discharge destinations | 11 |
| 6 | Counts of TBI codes | 12 |
| 7 | Counts of top NTT codes | 13 |
| 8 | Post-matching demographics of study population | 14 |
| 9 | Demographics of study population with and without dementia | 15 |
| 10 | Post-matching demographics of study population with and without dementia | 16 |
| 11 | Cox regression model for impact of injury on 1M survival | 17 |
| 12 | Cox regression results for impact of injury on 1M survival | 17 |
| 13 | Cox regression model for impact of injury on 6M survival | 18 |
| 14 | Cox regression results for impact of injury on 6M survival | 18 |
| 15 | Cox regression model for impact of injury on 12M survival | 19 |
| 16 | Cox regression results for impact of injury on 12M survival | 19 |
| 17 | Cox regression model for impact of injury and dementia on 1M survival | 20 |
| 18 | Cox regression results for impact of injury and dementia on 1M survival | 20 |
| 19 | Cox regression model for impact of injury and dementia on 6M survival | 21 |
| 20 | Cox regression results for impact of injury and dementia on 6M survival | 21 |
| 21 | Cox regression model for impact of injury and dementia on 12M survival | 22 |
| 22 | Cox regression results for impact of injury and dementia on 12M survival | 22 |

| **Figures** | | |
|---|---|---|
| 1 | Counts of mechanisms associated with NTT by age and sex | 22 |
| 2 | Counts of mechanisms associated with TBI by age and sex | 23 |
| 3 | Mechanisms associated with dementia trauma– post-matching | 24 |
| 4 | Mechanisms associated with non-dementia trauma – post matching | 25 |
| 5 | Impact of deprivation on survival at 1M | 26 |
| 6 | Impact of deprivation on survival at 6M | 27 |
| 7 | Impact of deprivation on survival at 12M | 28 |

| **Results** | | |
|---|---|---|
| 1 | Extended cohort description – study population | 29 |
| 2 | Extended cohort description – study population with and without dementia | 29 |
| 3 | Extended description of NTT injury mechanisms | 29 |
| 4 | Extended description of Cox regression results – impact of injury on survival | 29 |
| 5 | Extended description of Cox regression results – impact of injury and dementia on survival | 29-30 |

**Supplementary Table 1. ICD-10 Codes for traumatic brain injury (TBI)**

| Code | Description |
|---|---|
| S06 | Intracranial injury |
| S06.0 | Concussion |
| S06.1 | Traumatic cerebral oedema |
| S06.2 | Diffuse brain injury |
| S06.3 | Focal brain injury |
| S06.4 | Epidural haemorrhage |
| S06.5 | Traumatic subdural haemorrhage |
| S06.6 | Traumatic subarachnoid haemorrhage |
| S06.7 | Intracranial injury with prolonged coma |
| S06.8 | Other intracranial injuries |
| S06.9 | Intracranial injury, unspecified |

**Supplementary Table 2. ICD-10 Codes for non-TBI trauma (NTT)**

| Code | Description |
|---|---|
| S01 | Open wound of head |
| S02 | Fracture of skull and facial bones |
| S03 | Dislocation, sprain and strain of joints and ligaments of head |
| S04 | Injury of cranial nerves |
| S05 | Injury of eye and orbit |
| S07 | Crushing injury of head |
| S08 | Traumatic amputation of part of head |
| S09 | Other and unspecified injuries of head |
| S11 | Open wound of neck |
| S12 | Fracture of neck |
| S13 | Dislocation, sprain and strain of joints and ligaments at neck level |
| S14 | Injury of nerves and spinal cord at neck level |
| S15 | Injury of blood vessels at neck level |
| S16 | Injury of muscle and tendon at neck level |
| S17 | Crushing injury of neck |
| S18 | Traumatic amputation at neck level |
| S19 | Other and unspecified injuries of neck |
| S21 | Open wound of thorax |
| S22 | Fracture of rib(s), sternum and thoracic spine |
| S23 | Dislocation, sprain and strain of joints and ligaments of thorax |
| S24 | Injury of nerves and spinal cord at thorax level |
| S25 | Injury of blood vessels of thorax |
| S26 | Injury of heart |
| S27 | Injury of other and unspecified intrathoracic organs |
| S28 | Crushing injury of thorax and traumatic amputation of part of thorax |
| S29 | Other and unspecified injuries of thorax |
| S31 | Open wound of abdomen, lower back and pelvis |
| S32 | Fracture of lumbar spine and pelvis |
| S33 | Dislocation, sprain and strain of joints and ligaments of lumbar spine and pelvis |
| S34 | Injury of nerves and lumbar spinal cord at abdomen, lower back and pelvis level |
| S35 | Injury of blood vessels at abdomen, lower back and pelvis level |
| S36 | Injury of intra-abdominal organs |
| S37 | Injury of urinary and pelvic organs |
| S38 | Crushing injury and traumatic amputation of part of abdomen, lower back and pelvis |
| S39 | Other and unspecified injuries of abdomen, lower back and pelvis |
| S41 | Open wound of shoulder and upper arm |
| S42 | Fracture of shoulder and upper arm |
| S43 | Dislocation, sprain and strain of joints and ligaments of shoulder girdle |
| S44 | Injury of nerves at shoulder and upper arm level |
| S45 | Injury of blood vessels at shoulder and upper arm level |
| S46 | Injury of muscle and tendon at shoulder and upper arm level |
| S47 | Crushing injury of shoulder and upper arm |
| S48 | Traumatic amputation of shoulder and upper arm |
| S49 | Other and unspecified injuries of shoulder and upper arm |
| S51 | Open wound of forearm |
| S52 | Fracture of forearm |
| S53 | Dislocation, sprain and strain of joints and ligaments of elbow |

| | |
|---|---|
| S54 | Injury of nerves at forearm level |
| S55 | Injury of blood vessels at forearm level |
| S56 | Injury of muscle and tendon at forearm level |
| S57 | Crushing injury of forearm |
| S58 | Traumatic amputation of forearm |
| S59 | Other and unspecified injuries of forearm |
| S61 | Open wound of wrist and hand |
| S62 | Fracture at wrist and hand level |
| S63 | Dislocation, sprain and strain of joints and ligaments at wrist and hand level |
| S64 | Injury of nerves at wrist and hand level |
| S65 | Injury of blood vessels at wrist and hand level |
| S66 | Injury of muscle and tendon at wrist and hand level |
| S67 | Crushing injury of wrist and hand |
| S68 | Traumatic amputation of wrist and hand |
| S69 | Other and unspecified injuries of wrist and hand |
| S71 | Open wound of hip and thigh |
| S72 | Fracture of femur |
| S73 | Dislocation, sprain and strain of joint and ligaments of hip |
| S74 | Injury of nerves at hip and thigh level |
| S75 | Injury of blood vessels at hip and thigh level |
| S76 | Injury of muscle and tendon at hip and thigh level |
| S77 | Crushing injury of hip and thigh |
| S78 | Traumatic amputation of hip and thigh |
| S79 | Other and unspecified injuries of hip and thigh |
| S81 | Open wound of lower leg |
| S82 | Fracture of lower leg, including ankle |
| S83 | Dislocation, sprain and strain of joints and ligaments of knee |
| S84 | Injury of nerves at lower leg level |
| S85 | Injury of blood vessels at lower leg level |
| S86 | Injury of muscle and tendon at lower leg level |
| S87 | Crushing injury of lower leg |
| S88 | Traumatic amputation of lower leg |
| S89 | Other and unspecified injuries of lower leg |
| S91 | Open wound of ankle and foot |
| S92 | Fracture of foot, except ankle |
| S93 | Dislocation, sprain and strain of joints and ligaments at ankle and foot level |
| S94 | Injury of nerves at ankle and foot level |
| S95 | Injury of blood vessels at ankle and foot level |
| S96 | Injury of muscle and tendon at ankle and foot level |
| S97 | Crushing injury of ankle and foot |
| S98 | Traumatic amputation of ankle and foot |
| S99 | Other and unspecified injuries of ankle and foot |
| T07 | Unspecified multiple injuries |
| T14 | Injury of unspecified body region |

**Supplementary Table 3. ICD-10 and Read V2 codes for dementia**

| Read code | Description |
|---|---|
| A41.. | Slow viral central nervous system infection |
| A410. | Kuru |
| A411. | Creutzfeldt-Jakob disease |
| A413. | Progressive multifocal leucoencephalopathy |
| A41y. | Other slow virus central nervous system infections |
| A41z. | Slow virus central nervous system infection NOS |
| Ayu82 | [X]Other slow virus infections of central nervous system |
| Ayu83 | [X]Slow virus infection/central nervous system, unspecified |
| E000. | Uncomplicated senile dementia |
| E001. | Presenile dementia |
| E0010 | Uncomplicated presenile dementia |
| E0012 | Presenile dementia with paranoia |
| E0013 | Presenile dementia with depression |
| E002. | Senile dementia with depressive or paranoid features |
| E0020 | Senile dementia with paranoia |
| E0021 | Senile dementia with depression |
| E0040 | Uncomplicated arteriosclerotic dementia |
| E0041 | Arteriosclerotic dementia with delirium |
| E0042 | Arteriosclerotic dementia with paranoia |
| E0043 | Arteriosclerotic dementia with depression |
| Eu00. | [X]Dementia in Alzheimer's disease |
| Eu002 | [X]Dementia in Alzheimer's dis, atypical or mixed type |
| Eu00z | [X]Dementia in Alzheimer's disease, unspecified |
| Eu01y | [X]Other vascular dementia |
| Eu01z | [X]Vascular dementia, unspecified |
| Eu02. | [X]Dementia in other diseases classified elsewhere |
| Eu020 | [X]Dementia in Pick's disease |
| Eu021 | [X]Dementia in Creutzfeldt-Jakob disease |
| Eu022 | [X]Dementia in Huntington's disease |
| Eu023 | [X]Dementia in Parkinson's disease |
| Eu02y | [X]Dementia in other specified diseases classif elsewhere |
| F0301 | Subacute sclerosing panencephalitis |
| F1… | Hereditary and degenerative diseases of the CNS |
| F10y0 | Progressive neuronal degeneration with liver cirrhosis |
| F11.. | Other cerebral degenerations |
| F110. | Alzheimer's disease |
| F111. | Pick's disease |
| F112. | Senile degeneration of brain |
| F11x0 | Alcoholic encephalopathy |
| F11x7 | Cerebral degeneration due to Creutzfeldt-Jakob disease |
| F11x8 | Cerebral degeneration due to multifocal leucoencephalopathy |
| F11xz | Cerebral degeneration other disease NOS |
| F11yz | Other cerebral degeneration NOS |
| F1440 | Alcoholic cerebellar degeneration |
| Fyu30 | [X]Other Alzheimer's disease |
| Fyu31 | [X]Other specified degenerative diseases/the nervous system |
| X002w | Dementia |
| X002x | Dementia in Alzheimer's disease with early onset |
| X002y | Familial Alzheimer's disease of early onset |
| X002z | Non-familial Alzheimer's disease of early onset |

| | |
|---|---|
| X0030 | Dementia in Alzheimer's disease with late onset |
| X0031 | Familial Alzheimer's disease of late onset |
| X0032 | Non-familial Alzheimer's disease of late onset |
| X0033 | Focal Alzheimer's disease |
| X003R | Vascular dementia of acute onset |
| X003T | Subcortical vascular dementia |
| X003V | Mixed cortical and subcortical vascular dementia |
| X00R2 | Senile dementia |
| Xa01H | Multi-infarct dementia |
| Xa0sE | Dementia of frontal lobe type |
| XaIKB | Alzheimer's disease with early onset |
| XaIKC | Alzheimer's disease with late onset |
| XaIRJ | Cereb autosom dominant arteriop subcort infarcts leukoenceph |
| XE1Xs | Vascular dementia |
| XE1Z6 | [X]Unspecified dementia |

**Supplementary Table 4. ICD-10 Codes for injury mechanisms**

| Code | Description | Grouping |
|---|---|---|
| V01 | Pedestrian injured in collision with pedal cycle | RTC |
| V02 | Pedestrian injured in collision with two- or three-wheeled motor vehicle | RTC |
| V03 | Pedestrian injured in collision with car, pick-up truck or van | RTC |
| V04 | Pedestrian injured in collision with heavy transport vehicle or bus | RTC |
| V05 | Pedestrian injured in collision with railway train or railway vehicle | RTC |
| V06 | Pedestrian injured in collision with other nonmotor vehicle | RTC |
| V09 | Pedestrian injured in other and unspecified transport accidents | RTC |
| V10 | Pedal cyclist injured in collision with pedestrian or animal | RTC |
| V11 | Pedal cyclist injured in collision with other pedal cycle | RTC |
| V12 | Pedal cyclist injured in collision with two- or three-wheeled motor vehicle | RTC |
| V13 | Pedal cyclist injured in collision with car, pick-up truck or van | RTC |
| V14 | Pedal cyclist injured in collision with heavy transport vehicle or bus | RTC |
| V15 | Pedal cyclist injured in collision with railway train or railway vehicle | RTC |
| V16 | Pedal cyclist injured in collision with other nonmotor vehicle | RTC |
| V17 | Pedal cyclist injured in collision with fixed or stationary object | RTC |
| V18 | Pedal cyclist injured in noncollision transport accident | RTC |
| V19 | Pedal cyclist injured in other and unspecified transport accidents | RTC |
| V20 | Motorcycle rider injured in collision with pedestrian or animal | RTC |
| V21 | Motorcycle rider injured in collision with pedal cycle | RTC |
| V22 | Motorcycle rider injured in collision with two- or three-wheeled motor vehicle | RTC |
| V23 | Motorcycle rider injured in collision with car, pick-up truck or van | RTC |
| V24 | Motorcycle rider injured in collision with heavy transport vehicle or bus | RTC |
| V25 | Motorcycle rider injured in collision with railway train or railway vehicle | RTC |
| V26 | Motorcycle rider injured in collision with other nonmotor vehicle | RTC |
| V27 | Motorcycle rider injured in collision with fixed or stationary object | RTC |
| V28 | Motorcycle rider injured in noncollision transport accident | RTC |
| V29 | Motorcycle rider injured in other and unspecified transport accidents | RTC |

| | | |
|---|---|---|
| V30 | Motorcycle rider injured in other and unspecified transport accidents | RTC |
| V31 | Occupant of three-wheeled motor vehicle injured in collision with pedal cycle | RTC |
| V32 | Occupant of three-wheeled motor vehicle injured in collision with two- or three-wheeled motor vehicle | RTC |
| V33 | Occupant of three-wheeled motor vehicle injured in collision with car, pick-up truck or van | RTC |
| V34 | Occupant of three-wheeled motor vehicle injured in collision with heavy transport vehicle or bus | RTC |
| V35 | Occupant of three-wheeled motor vehicle injured in collision with railway train or railway vehicle | RTC |
| V36 | Occupant of three-wheeled motor vehicle injured in collision with other nonmotor vehicle | RTC |
| V37 | Occupant of three-wheeled motor vehicle injured in collision with fixed or stationary object | RTC |
| V38 | Occupant of three-wheeled motor vehicle injured in noncollision transport accident | RTC |
| V39 | Occupant of three-wheeled motor vehicle injured in other and unspecified transport accidents | RTC |
| V40 | Car occupant injured in collision with pedestrian or animal | RTC |
| V41 | Car occupant injured in collision with pedal cycle | RTC |
| V42 | Car occupant injured in collision with two- or three-wheeled motor vehicle | RTC |
| V43 | Car occupant injured in collision with car, pick-up truck or van | RTC |
| V44 | Car occupant injured in collision with heavy transport vehicle or bus | RTC |
| V45 | Car occupant injured in collision with railway train or railway vehicle | RTC |
| V46 | Car occupant injured in collision with other nonmotor vehicle | RTC |
| V47 | Car occupant injured in collision with fixed or stationary object | RTC |
| V48 | Car occupant injured in noncollision transport accident | RTC |
| V49 | Car occupant injured in other and unspecified transport accidents | RTC |
| V50 | Occupant of pick-up truck or van injured in collision with pedestrian or animal | RTC |
| V51 | Occupant of pick-up truck or van injured in collision with pedal cycle | RTC |
| V52 | Occupant of pick-up truck or van injured in collision with two- or three-wheeled motor vehicle | RTC |
| V53 | Occupant of pick-up truck or van injured in collision with car, pick-up truck or van | RTC |
| V54 | Occupant of pick-up truck or van injured in collision with heavy transport vehicle or bus | RTC |
| V55 | Occupant of pick-up truck or van injured in collision with railway train or railway vehicle | RTC |
| V56 | Occupant of pick-up truck or van injured in collision with other nonmotor vehicle | RTC |
| V57 | Occupant of pick-up truck or van injured in collision with fixed or stationary object | RTC |

| | | |
|---|---|---|
| V58 | Occupant of pick-up truck or van injured in noncollision transport accident | RTC |
| V59 | Occupant of pick-up truck or van injured in other and unspecified transport accidents | RTC |
| V60 | Occupant of heavy transport vehicle injured in collision with pedestrian or animal | RTC |
| V61 | Occupant of heavy transport vehicle injured in collision with pedal cycle | RTC |
| V62 | Occupant of heavy transport vehicle injured in collision with two- or three-wheeled motor vehicle | RTC |
| V63 | Occupant of heavy transport vehicle injured in collision with car, pick-up truck or van | RTC |
| V64 | Occupant of heavy transport vehicle injured in collision with heavy transport vehicle or bus | RTC |
| V65 | Occupant of heavy transport vehicle injured in collision with railway train or railway vehicle | RTC |
| V66 | Occupant of heavy transport vehicle injured in collision with other nonmotor vehicle | RTC |
| V67 | Occupant of heavy transport vehicle injured in collision with fixed or stationary object | RTC |
| V68 | Occupant of heavy transport vehicle injured in noncollision transport accident | RTC |
| V69 | Occupant of heavy transport vehicle injured in other and unspecified transport accidents | RTC |
| V70 | Bus occupant injured in collision with pedestrian or animal | RTC |
| V71 | Bus occupant injured in collision with pedal cycle | RTC |
| V72 | Bus occupant injured in collision with two- or three-wheeled motor vehicle | RTC |
| V73 | Bus occupant injured in collision with car, pick-up truck or van | RTC |
| V74 | Bus occupant injured in collision with heavy transport vehicle or bus | RTC |
| V75 | Bus occupant injured in collision with railway train or railway vehicle | RTC |
| V76 | Bus occupant injured in collision with other nonmotor vehicle | RTC |
| V77 | Bus occupant injured in collision with fixed or stationary object | RTC |
| V78 | Bus occupant injured in noncollision transport accident | RTC |
| V79 | Bus occupant injured in other and unspecified transport accidents | RTC |
| V80 | Animal-rider or occupant of animal-drawn vehicle injured in transport accident | RTC |
| V81 | Occupant of railway train or railway vehicle injured in transport accident | RTC |
| V82 | Occupant of streetcar injured in transport accident | RTC |
| V83 | Occupant of special vehicle mainly used on industrial premises injured in transport accident | RTC |
| V84 | Occupant of special vehicle mainly used in agriculture injured in transport accident | RTC |
| V85 | Occupant of special construction vehicle injured in transport accident | RTC |

| | | |
|---|---|---|
| V86 | Occupant of special all-terrain or other motor vehicle designed primarily for off-road use, injured in transport accident | RTC |
| V87 | Traffic accident of specified type but victim's mode of transport unknown | RTC |
| V88 | Nontraffic accident of specified type but victim's mode of transport unknown | RTC |
| V89 | Motor- or nonmotor-vehicle accident, type of vehicle unspecified | RTC |
| V98 | Other specified transport accidents | RTC |
| V99 | Unspecified transport accident | RTC |
| W00 | Fall on same level involving ice and snow | Fall |
| W01 | Fall on same level from slipping, tripping and stumbling | Fall |
| W02 | Fall involving ice-skates, skis, roller-skates or skateboards | Fall |
| W03 | Other fall on same level due to collision with, or pushing by, another person | Fall |
| W04 | Fall while being carried or supported by other persons | Fall |
| W05 | Fall involving wheelchair | Fall |
| W06 | Fall involving bed | Fall |
| W07 | Fall involving chair | Fall |
| W08 | Fall involving other furniture | Fall |
| W09 | Fall involving playground equipment | Fall |
| W10 | Fall on and from stairs and steps | Fall |
| W11 | Fall on and from ladder | Fall |
| W12 | Fall on and from scaffolding | Fall |
| W13 | Fall from, out of or through building or structure | Fall |
| W14 | Fall from tree | Fall |
| W15 | Fall from cliff | Fall |
| W17 | Other fall from one level to another | Fall |
| W18 | Other fall on same level | Fall |
| W19 | Unspecified fall | Fall |
| X93 | Assault by handgun discharge | Assault |
| X94 | Assault by rifle, shotgun and larger firearm discharge | Assault |
| X95 | Assault by other and unspecified firearm discharge | Assault |
| X96 | Assault by explosive material | Assault |
| X99 | Assault by sharp object | Assault |
| Y00 | Assault by blunt object | Assault |
| Y01 | Assault by pushing from high place | Assault |
| Y02 | Assault by pushing or placing victim before moving object | Assault |
| Y03 | Assault by crashing of motor vehicle | Assault |
| Y04 | Assault by bodily force | Assault |
| Y05 | Sexual assault by bodily force | Assault |
| Y07 | Other maltreatment | Assault |
| Y08 | Assault by other specified means | Assault |
| Y09 | Assault by unspecified means | Assault |
| Y87 | Sequelae of intentional self-harm, assault and events of undetermined intent | Assault |
| T76 | Unspecified effects of external causes | Assault |
| T74 | Maltreatment syndromes | Assault |

**Supplementary Table 5. Code mappings for discharge destinations**

| Value | Description (NHS Wales Data Dictionary) | Grouping |
|---|---|---|
| 19 | Own home | Leaving medical care |
| 20 | Permanent residence at nursing home, residential care home | Social care/care home care |
| 21 | Temporary residence at nursing home, residential care home | Social care/care home care |
| 22 | No fixed abode | Leaving medical care |
| 23 | Hospice | Continuing medical care |
| 29 | Temporary place of residence when usually resident elsewhere (includes hotel, residential educational establishment) | Leaving medical care |
| 39 | Penal establishment, court or police station or police custody suite | Leaving medical care |
| 49 | *Special Hospital* | Continuing medical care |
| 51 | Patient transfer to another health board / trust | Continuing medical care |
| 52 | Other Local Health Board/Trust - ward for maternity patients or neonates | Continuing medical care |
| 53 | Other Local Health Board/Trust - ward for patients who are mentally ill or have learning disabilities | Continuing medical care |
| 54 | NHS run nursing home, group home or residential care home | Social care/care home care |
| 55 | Patient transfer within the same health board / trust | Continuing medical care |
| 56 | Hospital site within the same Local Health Board/Trust - ward for maternity patients or neonates | Continuing medical care |
| 57 | Hospital site within the same Local Health Board/Trust - ward for patients who are mentally ill or have learning disabilities | Continuing medical care |
| 65 | Local Authority Part 3 residential accommodation i.e. where care is provided | Social care/care home care |
| 66 | Local authority foster care but not in Part 3 residential accommodation | Social care/care home care |
| 69 | *Under local authority care – residential or foster care* | Social care/care home care |
| 79 | Not Applicable - Patient died or stillbirth | Death |
| 85 | Non-NHS (other than local authority) run residential care home | Social care/care home care |
| 86 | Non-NHS (other than local authority) run nursing home | Social care/care home care |
| 87 | Patient transfer to Non-NHS run hospital includes private hospitals e.g. BUPA | Continuing medical care |
| 88 | Non-NHS (other than Local Authority) run Hospice | Continuing medical care |
| 89 | *Other non-NHS Hospital, Nursing Home or Residential institution* | Other/Unknown |
| 98 | Not applicable - hospital provider spell not finished at episode end (i.e. not discharged, or current episode unfinished) | Other/Unknown |
| 99 | *Not Known* | Other/Unknown |
| NS | *Hospital provider spell not yet finished* | Other/Unknown |

**Supplementary Table 6. Counts of TBI codes**

| ICD-10 | Description | Count |
|---|---|---|
| S065 | Traumatic subdural haemorrhage | 9607 |
| S062 | Diffuse brain injury | 2925 |
| S066 | Traumatic subarachnoid haemorrhage | 2817 |
| S068 | Other intracranial injuries | 2103 |
| S063 | Focal brain injury | 1674 |
| S060 | Concussion | 1459 |
| S069 | Intracranial injury unspecified | 1417 |
| S064 | Epidural haemorrhage | 812 |
| S061 | Traumatic cerebral oedema | 604 |
| S067 | Intracranial injury with prolonged coma | 10 |

**Supplementary Table 7. Counts of top NTT codes**

| ICD-10 | Description | Count |
|---|---|---|
| S72 | Fracture of neck of femur | 107322 |
| S82 | Fracture of lower leg, including ankle | 54272 |
| S01 | Open wound of head | 50235 |
| S61 | Open wound of wrist and hand | 46728 |
| S52 | Fracture of forearm | 42910 |
| S42 | Fracture of shoulder and upper arm | 31062 |
| S32 | Fracture of lumbar spine and pelvis | 28358 |
| S09 | Other and unspecified injuries of head | 28053 |
| S02 | Fracture of skull and facial bones | 27839 |
| S22 | Fracture of rib(s), sternum and thoracic spine | 25801 |
| S62 | Fracture at wrist and hand level | 21850 |
| S83 | Dislocation, sprain and strain of joints and ligaments of knee | 11705 |
| S51 | Open wound of forearm | 11443 |
| S81 | Open wound of lower leg | 10239 |
| S92 | Fracture of foot, except ankle | 8789 |
| S43 | Dislocation, sprain and strain of joints and ligaments of shoulder girdle | 5846 |
| S12 | Fracture of neck | 4380 |
| S66 | Injury of muscle and tendon at wrist and hand level | 4334 |
| S39 | Other and unspecified injuries of abdomen, lower back and pelvis | 3758 |
| S05 | Injury of eye and orbit | 3736 |
| S91 | Open wound of ankle and foot | 3678 |
| S86 | Injury of muscle and tendon at lower leg level | 3617 |
| S31 | Open wound of abdomen, lower back and pelvis | 3514 |
| S68 | Traumatic amputation of wrist and hand | 3057 |
| S46 | Injury of muscle and tendon at shoulder and upper arm level | 2968 |
| S63 | Dislocation, sprain and strain of joints and ligaments at wrist and hand level | 2922 |
| S79 | Other and unspecified injuries of hip and thigh | 2642 |
| T14 | Injury of unspecified body region | 2527 |
| S76 | Injury of muscle and tendon at hip and thigh level | 2420 |
| S41 | Open wound of shoulder and upper arm | 2214 |

### Supplementary Table 8. Post-matching demographics of study population

| | | Overall | Non-TBI | TBI |
|---|---|---|---|---|
| **n** | | 85881 | 72481 | 13400 |
| **Age, mean (SD)** | | 71.6 (15.9) | 71.7 (15.8) | 71.4 (16.0) |
| **Sex, n (%)** | **Female** | 36796 (42.8) | 31102 (42.9) | 5694 (42.5) |
| | **Male** | 49085 (57.2) | 41379 (57.1) | 7706 (57.5) |
| **Region, n (%)** | **Rural** | 28972 (33.7) | 24117 (33.3) | 4855 (36.2) |
| | **Urban** | 56909 (66.3) | 48364 (66.7) | 8545 (63.8) |
| **WIMD Quintile (2019), n (%)** | **1 (Most deprived)** | 17259 (20.1) | 14594 (20.1) | 2665 (19.9) |
| | **2** | 17970 (20.9) | 15232 (21.0) | 2738 (20.4) |
| | **3** | 17716 (20.6) | 14945 (20.6) | 2771 (20.7) |
| | **4** | 17077 (19.9) | 14397 (19.9) | 2680 (20.0) |
| | **5 (Least deprived)** | 15857 (18.5) | 13311 (18.4) | 2546 (19.0) |
| **Mechanism of injury, n (%)** | **Assault** | 1501 (1.7) | 1158 (1.6) | 343 (2.6) |
| | **Fall** | 56134 (65.4) | 47273 (65.2) | 8861 (66.1) |
| | **Other/Unknown** | 23691 (27.6) | 20452 (28.2) | 3239 (24.2) |
| | **RTC** | 4555 (5.3) | 3598 (5.0) | 957 (7.1) |
| **Discharge Destination, n (%)** | **Death** | 6326 (7.4) | 4165 (5.8) | 2161 (16.1) |
| | **Leaving medical care** | 62016 (72.3) | 54203 (74.8) | 7813 (58.4) |
| | **Ongoing medical care** | 14756 (17.2) | 11794 (16.3) | 2962 (22.1) |
| | **Other/Unknown** | 82 (0.1) | 61 (0.1) | 21 (0.2) |
| | **Social/carehome care** | 2627 (3.1) | 2196 (3.0) | 431 (3.2) |
| **Comorbidities, n (%)** | **Psychiatric** | 20171 (23.5) | 17070 (23.6) | 3101 (23.1) |
| | **Neurodegenerative (non-dementia)** | 1690 (2.0) | 1496 (2.1) | 194 (1.4) |
| | **Diabetes** | 23312 (27.1) | 19744 (27.2) | 3568 (26.6) |
| | **Cancer** | 33362 (38.8) | 28212 (38.9) | 5150 (38.4) |
| | **Respiratory** | 9940 (11.6) | 8490 (11.7) | 1450 (10.8) |
| | **Cardiovascular** | 55680 (64.8) | 46972 (64.8) | 8708 (65.0) |
| | **Liver** | 3558 (4.1) | 2986 (4.1) | 572 (4.3) |
| | **Renal** | 3774 (4.4) | 3155 (4.4) | 619 (4.6) |
| | **Cerebrovascular** | 33079 (38.5) | 27836 (38.4) | 5243 (39.1) |

### Supplementary Table 9. Demographics of study population with and without dementia

| | | Overall | DEM NTT | DEM TBI | ND NTT | ND TBI |
|---|---|---|---|---|---|---|
| **Episodes** | | 449,047 | 59,785 | 3,083 | 370,751 | 15,428 |
| **N** | | 311,230 | 39,852 | 2,460 | 266,082 | 12,420 |
| **Age, mean (SD)** | | 71.4 (15.6) | 83.1 (9.3) | 81.5 (10.9) | 69.4 (15.7) | 69.9 (15.2) |
| **Sex, n (%)** | **Female** | 263,249 (58.6) | 41,350 (69.2) | 1,630 (52.9) | 213,818 (57.7) | 6,451 (41.8) |
| | **Male** | 185,798 (41.4) | 18,435 (30.8) | 1,453 (47.1) | 156,933 (42.3) | 8,977 (58.2) |
| **Region, n (%)** | **Rural** | 134,050 (31.4) | 15,713 (28.2) | 798 (27.8) | 112,911 (31.9) | 4,628 (31.5) |
| | **Urban** | 293,504 (68.6) | 40,000 (71.8) | 2,077 (72.2) | 241,356 (68.1) | 10,071 (68.5) |
| **Welsh Index of Multiple Deprivation (WIMD) Quintile (2019), n (%)** | **1 (Most deprived)** | 91,899 (20.5) | 11,609 (19.4) | 607 (19.7) | 76,494 (20.6) | 3,189 (20.7) |
| | **2** | 95,736 (21.3) | 12,972 (21.7) | 652 (21.1) | 78,944 (21.3) | 3,168 (20.5) |
| | **3** | 92,769 (20.7) | 11,845 (19.8) | 634 (20.6) | 77,194 (20.8) | 3,096 (20.1) |
| | **4** | 87,829 (19.6) | 11,767 (19.7) | 592 (19.2) | 72,372 (19.5) | 3,098 (20.1) |
| | **5 (Least deprived)** | 80,772 (18.0) | 11,592 (19.4) | 598 (19.4) | 65,705 (17.7) | 2,877 (18.6) |
| **Mechanism of injury, n (%)** | **Assault** | 7,150 (1.6) | 178 (0.3) | 20 (0.6) | 6,540 (1.8) | 412 (2.7) |
| | **Fall** | 265,629 (59.2) | 41,521 (69.5) | 2,161 (70.1) | 212,966 (57.4) | 8,981 (58.2) |
| | **Other/Unknown** | 155,645 (34.7) | 17,682 (29.6) | 874 (28.3) | 132,136 (35.6) | 4,953 (32.1) |
| | **Road traffic collision (RTC)** | 20,623 (4.6) | 404 (0.7) | 28 (0.9) | 19,109 (5.2) | 1,082 (7.0) |
| **Discharge Destination, n (%)** | **Death/ Other /Unknown** | 25,908 (5.77) | 6,524 (10.9) | 610 (19.8) | 16,513 (4.45) | 2,261 (14.7) |
| | **Leaving medical care** | 329,979 (73.6) | 29,636 (49.7) | 1,399 (45.4) | 289,714 (78.3) | 9,230 (59.9) |
| | **Ongoing medical care** | 73,875 (16.5) | 14,786 (24.8) | 708 (23.0) | 54,887 (14.8) | 3,494 (22.7) |
| | **Social/ Care home** | 18,377 (4.1) | 8,685 (14.6) | 364 (11.8) | 8,909 (2.4) | 419 (2.7) |
| **Comorbidities, n (%)** | **Psychiatric** | 109,169 (24.3) | 16,922 (28.3) | 1,038 (33.7) | 87,151 (23.5) | 4,058 (26.3) |
| | **Neurodegenerative (Non-dementia)** | 11,997 (2.7) | 3,468 (5.8) | 167 (5.4) | 8,044 (2.2) | 318 (2.1) |
| | **Diabetes** | 114,794 (25.6) | 21,792 (36.5) | 1,250 (40.5) | 87,540 (23.6) | 4,212 (27.3) |
| | **Cancer** | 168,600 (37.5) | 27,579 (46.1) | 1,494 (48.5) | 133,695 (36.1) | 5,832 (37.8) |
| | **Respiratory** | 66,274 (14.8) | 10,899 (18.2) | 509 (16.5) | 52,996 (14.3) | 1,870 (12.1) |
| | **Cardiovascular** | 280,098 (62.4) | 43,442 (72.7) | 2,311 (75.0) | 224,417 (60.5) | 9,928 (64.4) |
| | **Liver** | 16,793 (3.7) | 2,656 (4.4) | 214 (6.9) | 13,054 (3.5) | 869 (5.6) |
| | **Renal** | 19,673 (4.4) | 4,308 (7.2) | 229 (7.4) | 14,412 (3.9) | 724 (4.7) |
| | **Cerebrovascular** | 146,320 (32.6) | 30,480 (51.0) | 1,818 (59.0) | 108,100 (29.2) | 5,922 (38.4) |

### Supplementary Table 10. Post-matching demographics of study population with and without dementia

| | | Overall | DEM NTT | DEM TBI | ND NTT | ND TBI |
|---|---|---|---|---|---|---|
| **n** | | 42341 | 15188 | 3127 | 18659 | 5367 |
| **Age, mean (SD)** | | 81.4 (12.1) | 81.8 (12.2) | 81.4 (12.2) | 81.5 (11.9) | 80.4 (12.2) |
| **Sex, n (%)** | **Female** | 22436 (53.0) | 8179 (53.9) | 1646 (52.6) | 9853 (52.8) | 2758 (51.4) |
| | **Male** | 19905 (47.0) | 7009 (46.1) | 1481 (47.4) | 8806 (47.2) | 2609 (48.6) |
| **Region, n (%)** | **Rural** | 12097 (30.3) | 3946 (28.1) | 805 (27.6) | 5703 (32.0) | 1643 (32.0) |
| | **Urban** | 27813 (69.7) | 10111 (71.9) | 2107 (72.4) | 12101 (68.0) | 3494 (68.0) |
| **WIMD Quintile (2019), n (%)** | **1.0** | 8031 (19.0) | 2993 (19.7) | 622 (19.9) | 3469 (18.6) | 947 (17.6) |
| | **2.0** | 9056 (21.4) | 3306 (21.8) | 658 (21.0) | 3988 (21.4) | 1104 (20.6) |
| | **3.0** | 8697 (20.5) | 3007 (19.8) | 645 (20.6) | 3931 (21.1) | 1114 (20.8) |
| | **4.0** | 8460 (20.0) | 2960 (19.5) | 598 (19.1) | 3765 (20.2) | 1137 (21.2) |
| | **5.0** | 8097 (19.1) | 2922 (19.2) | 604 (19.3) | 3506 (18.8) | 1065 (19.8) |
| **Mechanism of injury, n (%)** | **Assault** | 223 (0.5) | 74 (0.5) | 26 (0.8) | 84 (0.5) | 39 (0.7) |
| | **Fall** | 28578 (67.5) | 10543 (69.4) | 2183 (69.8) | 12236 (65.6) | 3616 (67.4) |
| | **Other/Unknown** | 12682 (30.0) | 4427 (29.1) | 889 (28.4) | 5845 (31.3) | 1521 (28.3) |
| | **RTC** | 858 (2.0) | 144 (0.9) | 29 (0.9) | 494 (2.6) | 191 (3.6) |
| **Discharge Destination, n (%)** | **Death/Other/Unknown** | 4772 (11.3) | 1677 (11.0) | 613 (19.6) | 1471(7.9) | 1011 (18.8) |
| | **Leaving medical care** | 24253 (57.3) | 7865 (51.8) | 1431 (45.8) | 12121 (65.0) | 2836 (52.9) |
| | **Ongoing medical care** | 9868 (23.3) | 3611 (23.8) | 717 (22.9) | 4242 (22.8) | 1298 (24.2) |
| | **Social/carehome care** | 3403 (8.0) | 2017 (13.3) | 364 (11.6) | 808 (4.3) | 214 (4.0) |
| **Comorbidities, n (%)** | **Psychiatric** | 14023 (33.1) | 5024 (33.1) | 1060 (33.9) | 6306 (33.8) | 1633 (30.4) |
| | **Neurodegenerative (non-dementia)** | 1899 (4.5) | 698 (4.6) | 169 (5.4) | 911 (4.9) | 121 (2.3) |
| | **Diabetes** | 16651 (39.3) | 5891 (38.8) | 1254 (40.1) | 7461 (40.0) | 2045 (38.1) |
| | **Cancer** | 20426 (48.2) | 7375 (48.6) | 1509 (48.3) | 9035 (48.4) | 2507 (46.7) |
| | **Respiratory** | 6723 (15.9) | 2440 (16.1) | 512 (16.4) | 3004 (16.1) | 767 (14.3) |
| | **Cardiovascular** | 31651 (74.8) | 11350 (74.7) | 2328 (74.4) | 13922 (74.6) | 4051 (75.5) |
| | **Liver** | 2620 (6.2) | 973 (6.4) | 229 (7.3) | 1075 (5.8) | 343 (6.4) |
| | **Renal** | 2684 (6.3) | 1050 (6.9) | 229 (7.3) | 1067 (5.7) | 338 (6.3) |
| | **Cerebrovascular** | 24540 (58.0) | 8795 (57.9) | 1826 (58.4) | 10896 (58.4) | 3023 (56.3) |

### Supplementary Table 11. Cox regression model for impact of injury on 1M survival

| formula | observations | events | censored | log_likelihood | concordance | AIC | t_stats | df | p | log_p |
|---|---|---|---|---|---|---|---|---|---|---|
| SEX + C(AGE_BINS) + Injury + WIMD_2019_QUINTILE | 249593 | 12528 | 237065 | -149844.78 | 0.76 | 299707.57 | 11056.48 | 9 | 0 | inf |

### Supplementary Table 12. Cox regression results for impact of injury on 1M survival

| covariate | coef | exp(coef) | se(coef) | coef lower 95% | coef upper 95% | exp(coef) lower 95% | exp(coef) upper 95% | cmp_to | z | p | -log2(p) |
|---|---|---|---|---|---|---|---|---|---|---|---|
| SEX[T.Male] | 0.37212743 | 1.45081785 | 0.0183224 | 0.33621619 | 0.40803868 | 1.39964158 | 1.50386532 | 0 | 20.3099737 | 1.0496E-91 | 302.225609 |
| C(AGE_BINS)[T.40-64] | 0.57931549 | 1.78481629 | 0.04706377 | 0.48707219 | 0.67155879 | 1.6275441 | 1.95728594 | 0 | 12.3091597 | 8.0861E-35 | 113.252046 |
| C(AGE_BINS)[T.65-79] | 1.72941705 | 5.63736667 | 0.04200604 | 1.64708673 | 1.81174737 | 5.19183258 | 6.121134 | 0 | 41.1706782 | 0 | inf |
| C(AGE_BINS)[T.80-100] | 2.44464404 | 11.5264459 | 0.040073 | 2.36610239 | 2.52318568 | 10.6557792 | 12.4682533 | 0 | 61.0047621 | 0 | inf |
| Injury[T.TBI] | 1.19511143 | 3.3039259 | 0.02085318 | 1.15423995 | 1.23598291 | 3.17161191 | 3.4417598 | 0 | 57.3107523 | 0 | inf |
| WIMD_2019_QUINTILE[T.2.0] | -0.04196491 | 0.95890343 | 0.02680355 | -0.09449891 | 0.01056909 | 0.90982873 | 1.01062514 | 0 | -1.56564717 | 0.11743124 | 3.09011182 |
| WIMD_2019_QUINTILE[T.3.0] | -0.13879508 | 0.87040638 | 0.02755097 | -0.19279398 | -0.08479618 | 0.82465185 | 0.91869952 | 0 | -5.03775736 | 4.7102E-07 | 21.0177145 |
| WIMD_2019_QUINTILE[T.4.0] | -0.11896725 | 0.88783688 | 0.02763922 | -0.17313911 | -0.06479538 | 0.84102061 | 0.93725923 | 0 | -4.3042916 | 1.6752E-05 | 15.8652994 |
| WIMD_2019_QUINTILE[T.5.0] | -0.10527543 | 0.90007658 | 0.02795097 | -0.16005831 | -0.05049254 | 0.8520941 | 0.95076102 | 0 | -3.7664326 | 0.0001656 | 12.560379 |

### Supplementary Table 13. Cox regression model for impact of injury on 6M survival

| formula | observations | events | censored | log_likelihood | concordance | AIC | t_stats | df | p | log_p |
|---|---|---|---|---|---|---|---|---|---|---|
| SEX + C(AGE_BINS) + Injury + WIMD_2019_QUINTILE | 249593 | 31634 | 217959 | -376132.73 | 0.77 | 752283.45 | 29812.87 | 9 | 0 | inf |

### Supplementary Table 14. Cox regression results for impact of injury on 6M survival

| covariate | coef | exp(coef) | se(coef) | coef lower 95% | coef upper 95% | exp(coef) lower 95% | exp(coef) upper 95% | cmp to | z | p | -log2(p) |
|---|---|---|---|---|---|---|---|---|---|---|---|
| SEX[T.Male] | 0.348712 | 1.417241 | 0.011491 | 0.32619 | 0.371235 | 1.385678 | 1.449524 | 0 | 30.34537 | 2.9E-202 | 669.4972 |
| C(AGE_BINS)[T.40-64] | 0.906864 | 2.476543 | 0.03809 | 0.832208 | 0.981519 | 2.298389 | 2.668506 | 0 | 23.80836 | 2.7E-125 | 413.7887 |
| C(AGE_BINS)[T.65-79] | 2.288826 | 9.863354 | 0.034457 | 2.221292 | 2.35636 | 9.219235 | 10.55248 | 0 | 66.42583 | 0 | inf |
| C(AGE_BINS)[T.80-100] | 3.137223 | 23.0398 | 0.033426 | 3.071709 | 3.202737 | 21.57875 | 24.59977 | 0 | 93.85514 | 0 | inf |
| Injury[T.TBI] | 0.694612 | 2.002931 | 0.015517 | 0.6642 | 0.725024 | 1.942935 | 2.06478 | 0 | 44.76562 | 0 | inf |
| WIMD_2019_QUINTILE[T.2.0] | -0.06294 | 0.938998 | 0.017175 | -0.0966 | -0.02928 | 0.907916 | 0.971145 | 0 | -3.66473 | 0.000248 | 11.97971 |
| WIMD_2019_QUINTILE[T.3.0] | -0.1237 | 0.883642 | 0.017436 | -0.15788 | -0.08953 | 0.853955 | 0.914362 | 0 | -7.09468 | 1.3E-12 | 39.48854 |
| WIMD_2019_QUINTILE[T.4.0] | -0.16654 | 0.846591 | 0.017771 | -0.20137 | -0.13171 | 0.817611 | 0.876597 | 0 | -9.3714 | 7.16E-21 | 66.92103 |
| WIMD_2019_QUINTILE[T.5.0] | -0.14611 | 0.864064 | 0.017914 | -0.18122 | -0.111 | 0.834252 | 0.894941 | 0 | -8.15609 | 3.46E-16 | 51.35986 |

### Supplementary Table 15. Cox regression model for impact of injury on 12M survival

| formula | observations | events | censored | log_likelihood | concordance | AIC | t_stats | df | p | log_p |
|---|---|---|---|---|---|---|---|---|---|---|
| SEX + C(AGE_BINS) + Injury + WIMD_2019_QUINTILE | 249593 | 42343 | 207250 | -502132.06 | 0.77 | 1004282.11 | 40550.65 | 9 | 0 | inf |

### Supplementary Table 16. Cox regression results for impact of injury on 12M survival

| covariate | coef | exp(coef) | se(coef) | coef lower 95% | coef upper 95% | exp(coef) lower 95% | exp(coef) upper 95% | cmp to | z | p | -log2(p) |
|---|---|---|---|---|---|---|---|---|---|---|---|
| SEX[T.Male] | 0.335547 | 1.398706 | 0.00993 | 0.316085 | 0.35501 | 1.371746 | 1.426195 | 0 | 33.79077 | 2.7E-250 | 829.0516 |
| C(AGE_BINS)[T.40-64] | 1.054049 | 2.869246 | 0.034394 | 0.986637 | 1.121461 | 2.6822 | 3.069336 | 0 | 30.64593 | 3E-206 | 682.7349 |
| C(AGE_BINS)[T.65-79] | 2.417074 | 11.213 | 0.031524 | 2.355289 | 2.478859 | 10.54117 | 11.92765 | 0 | 76.67496 | 0 | inf |
| C(AGE_BINS)[T.80-100] | 3.29121 | 26.87538 | 0.030687 | 3.231066 | 3.351355 | 25.30661 | 28.54139 | 0 | 107.2519 | 0 | inf |
| Injury[T.TBI] | 0.56599 | 1.76119 | 0.014094 | 0.538366 | 0.593613 | 1.713205 | 1.810518 | 0 | 40.15857 | 0 | inf |
| WIMD_2019_QUINTILE[T.2.0] | -0.06271 | 0.939213 | 0.01487 | -0.09186 | -0.03357 | 0.912235 | 0.96699 | 0 | -4.21732 | 2.47E-05 | 15.30385 |
| WIMD_2019_QUINTILE[T.3.0] | -0.12043 | 0.886538 | 0.015076 | -0.14998 | -0.09088 | 0.860726 | 0.913124 | 0 | -7.98846 | 1.37E-15 | 49.37859 |
| WIMD_2019_QUINTILE[T.4.0] | -0.1806 | 0.834768 | 0.015428 | -0.21084 | -0.15036 | 0.809905 | 0.860395 | 0 | -11.7063 | 1.18E-31 | 102.7376 |
| WIMD_2019_QUINTILE[T.5.0] | -0.15715 | 0.854578 | 0.015545 | -0.18762 | -0.12668 | 0.828934 | 0.881016 | 0 | -10.1091 | 5.03E-24 | 77.39458 |

**Supplementary Table 17. Cox regression model for impact of injury and dementia on 1M survival**

| formula | observations | events | censored | log_likelihood | concordance | AIC | Ratio_test | t_stats | df | p | log_p |
|---|---|---|---|---|---|---|---|---|---|---|---|
| SEX + C(AGE_BINS) + C(WIMD_2019_QUINTILE) + GROUPS | 41724 | 4285 | 37439 | -44801.5 | 0.64 | 89623 | | 1115.94 | 10 | 1.9E-233 | 773.06 |

**Supplementary Table 18. Cox regression results for impact of injury and dementia on 1M survival**

| covariate | coef | exp(coef) | se(coef) | coef lower 95% | coef upper 95% | exp(coef) lower 95% | exp(coef) upper 95% | z | p | -log2(p) |
|---|---|---|---|---|---|---|---|---|---|---|
| SEX[T.Male] | 0.2859 | 1.33096 | 0.030973 | 0.225194 | 0.346606 | 1.252566 | 1.414259 | 9.23066 | 2.69E-20 | 65.01111 |
| C(AGE_BINS)[T.65-79] | 0.608242 | 1.837198 | 0.087668 | 0.436416 | 0.780067 | 1.547152 | 2.181618 | 6.938037 | 3.98E-12 | 37.87187 |
| C(AGE_BINS)[T.80-100] | 1.078887 | 2.941405 | 0.082593 | 0.917008 | 1.240767 | 2.501793 | 3.458265 | 13.06268 | 5.38E-39 | 127.1275 |
| C(WIMD_2019_QUINTILE)[T.2.0] | -0.00148 | 0.99852 | 0.048123 | -0.0958 | 0.092838 | 0.908644 | 1.097284 | -0.03079 | 0.97544 | 0.035875 |
| C(WIMD_2019_QUINTILE)[T.3.0] | -0.03337 | 0.967182 | 0.048719 | -0.12886 | 0.06212 | 0.8791 | 1.06409 | -0.68491 | 0.493398 | 1.019176 |
| C(WIMD_2019_QUINTILE)[T.4.0] | -0.08816 | 0.915613 | 0.049441 | -0.18507 | 0.008742 | 0.83105 | 1.00878 | -1.78316 | 0.074561 | 3.745436 |
| C(WIMD_2019_QUINTILE)[T.5.0] | -0.07351 | 0.929125 | 0.049662 | -0.17085 | 0.023825 | 0.842949 | 1.024111 | -1.48023 | 0.138812 | 2.848799 |
| GROUPS[T.DEM_NTT] | 0.248122 | 1.281616 | 0.038211 | 0.17323 | 0.323013 | 1.18914 | 1.381284 | 6.49353 | 8.38E-11 | 33.47343 |
| GROUPS[T.ND_TBI] | 1.05478 | 2.871344 | 0.041983 | 0.972495 | 1.137066 | 2.644534 | 3.117607 | 25.1239 | 2.7E-139 | 460.301 |
| GROUPS[T.DEM_TBI] | 0.982451 | 2.670996 | 0.050439 | 0.883592 | 1.081311 | 2.419575 | 2.948542 | 19.47783 | 1.69E-84 | 278.2824 |

**Supplementary Table 19. Cox regression results for impact of injury and dementia on 6M survival**

| formula | observations | events | censored | log_likelihood | concordance | AIC | Ratio_test | t_stats | df | p | log_p |
|---|---|---|---|---|---|---|---|---|---|---|---|
| SEX + C(AGE_BINS) + C(WIMD_2019_QUINTILE) + GROUPS | 41724 | 11013 | 30711 | -114490 | 0.62 | 228999.3 | | 2149.07 | 10 | 0 | inf |

**Supplementary Table 20. Cox regression results for impact of injury and dementia on 6M survival**

| covariate | coef | exp(coef) | se(coef) | coef lower 95% | coef upper 95% | exp(coef) lower 95% | exp(coef) upper 95% | z | p | -log2(p) |
|---|---|---|---|---|---|---|---|---|---|---|
| SEX[T.Male] | 0.307713 | 1.36031 | 0.019317 | 0.269852 | 0.345574 | 1.30977 | 1.4128 | 15.92945 | 3.96E-57 | 187.3652 |
| C(AGE_BINS)[T.65-79] | 0.748676 | 2.1142 | 0.059144 | 0.632757 | 0.864596 | 1.882795 | 2.374046 | 12.65863 | 1E-36 | 119.586 |
| C(AGE_BINS)[T.80-100] | 1.31709 | 3.732543 | 0.055991 | 1.207349 | 1.42683 | 3.344608 | 4.165475 | 23.52323 | 2.4E-122 | 404.0365 |
| C(WIMD_2019_QUINTILE)[T.2.0] | -0.02028 | 0.979923 | 0.029956 | -0.07899 | 0.038432 | 0.924046 | 1.03918 | -0.67702 | 0.498394 | 1.004642 |
| C(WIMD_2019_QUINTILE)[T.3.0] | -0.07274 | 0.929846 | 0.03047 | -0.13246 | -0.01302 | 0.875941 | 0.987069 | -2.38711 | 0.016982 | 5.87988 |
| C(WIMD_2019_QUINTILE)[T.4.0] | -0.11905 | 0.887765 | 0.030867 | -0.17955 | -0.05855 | 0.835649 | 0.943131 | -3.85681 | 0.000115 | 13.08763 |
| C(WIMD_2019_QUINTILE)[T.5.0] | -0.06045 | 0.941339 | 0.030693 | -0.12061 | -0.00029 | 0.88638 | 0.999706 | -1.96953 | 0.048892 | 4.35425 |
| GROUPS[T.DEM_NTT] | 0.448982 | 1.566716 | 0.022238 | 0.405395 | 0.492568 | 1.499895 | 1.636514 | 20.18946 | 1.21E-90 | 298.6965 |
| GROUPS[T.ND_TBI] | 0.583392 | 1.792106 | 0.029862 | 0.524862 | 0.641921 | 1.690226 | 1.900127 | 19.53602 | 5.43E-85 | 279.9242 |
| GROUPS[T.DEM_TBI] | 0.826416 | 2.285115 | 0.033232 | 0.761283 | 0.89155 | 2.141021 | 2.438907 | 24.86802 | 1.7E-136 | 451.059 |

**Supplementary Table 21. Cox regression results for impact of injury and dementia on 12M survival**

| formula | observations | events | censored | log_likelihood | concordance | AIC | Ratio_test | t_stats | df | p | log_p |
|---|---|---|---|---|---|---|---|---|---|---|---|
| SEX + C(AGE_BINS) + C(WIMD_2019_QUINTILE) + GROUPS | 41724 | 14553 | 27171 | -150551 | 0.62 | 301122 | | 2755.01 | 10 | 0 | inf |

**Supplementary Table 22. Cox regression results for impact of injury and dementia on 12M survival**

| covariate | coef | exp(coef) | se(coef) | coef lower 95% | coef upper 95% | exp(coef) lower 95% | exp(coef) upper 95% | z | p | -log2(p) |
|---|---|---|---|---|---|---|---|---|---|---|
| SEX[T.Male] | 0.311308 | 1.36521 | 0.016816 | 0.278349 | 0.344267 | 1.320947 | 1.410955 | 18.5126 | 1.63E-76 | 251.758 |
| C(AGE_BINS)[T.65-79] | 0.720595 | 2.055657 | 0.050459 | 0.621697 | 0.819493 | 1.862086 | 2.26935 | 14.28077 | 2.88E-46 | 151.2806 |
| C(AGE_BINS)[T.80-100] | 1.311298 | 3.710988 | 0.047677 | 1.217854 | 1.404743 | 3.379925 | 4.074478 | 27.50397 | 1.6E-166 | 550.786 |
| C(WIMD_2019_QUINTILE)[T.2.0] | -0.01454 | 0.985567 | 0.02605 | -0.0656 | 0.036519 | 0.93651 | 1.037194 | -0.55808 | 0.576787 | 0.79389 |
| C(WIMD_2019_QUINTILE)[T.3.0] | -0.06189 | 0.939984 | 0.026456 | -0.11375 | -0.01004 | 0.892485 | 0.99001 | -2.33948 | 0.019311 | 5.694455 |
| C(WIMD_2019_QUINTILE)[T.4.0] | -0.12681 | 0.880901 | 0.026907 | -0.17955 | -0.07407 | 0.835649 | 0.928602 | -4.71299 | 2.44E-06 | 18.64408 |
| C(WIMD_2019_QUINTILE)[T.5.0] | -0.06539 | 0.936705 | 0.026767 | -0.11785 | -0.01292 | 0.88883 | 0.987159 | -2.44278 | 0.014575 | 6.100396 |
| GROUPS[T.DEM_NTT] | 0.46035 | 1.584628 | 0.019028 | 0.423056 | 0.497643 | 1.526619 | 1.64484 | 24.19347 | 2.6E-129 | 427.1467 |
| GROUPS[T.ND_TBI] | 0.41819 | 1.519209 | 0.027006 | 0.365259 | 0.471121 | 1.440887 | 1.601789 | 15.48502 | 4.38E-54 | 177.2533 |
| GROUPS[T.DEM_TBI] | 0.764666 | 2.148278 | 0.029451 | 0.706944 | 0.822389 | 2.027785 | 2.27593 | 25.96431 | 1.3E-148 | 491.3193 |

**Supplementary Figure 1. Counts of mechanisms associated with NTT by age and sex.** *Males and females are mirrored, with males on the left and females on the right. Y-axis depicts age bands, with 20-year increments to prevent re-identification, with the youngest group (18-19) at the bottom and the oldest group (80-100) at the top; X-axis depicts counts of NTT episodes that were associated with the following mechanisms: Falls (Indigo), road traffic accidents (RTAs; dark purple), assault (light purple), and all other or unknown causes (pink).*

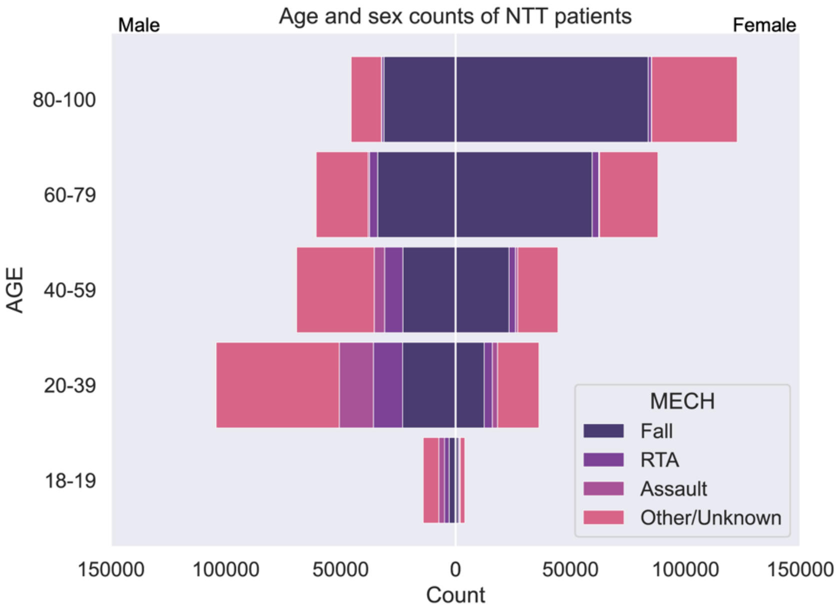

**Supplementary Figure 2. Counts of mechanisms associated with TBI by age and sex.** *Males and females are mirrored, with males on the left and females on the right. Y-axis depicts age bands, with 20-year increments to prevent re-identification, with the youngest group (18-19) at the bottom and the oldest group (80-100) at the top; X-axis depicts counts of TBI episodes that were associated with the following mechanisms: Falls (Indigo), road traffic accidents (RTAs; dark purple), assault (light purple), and all other or unknown causes (pink).*

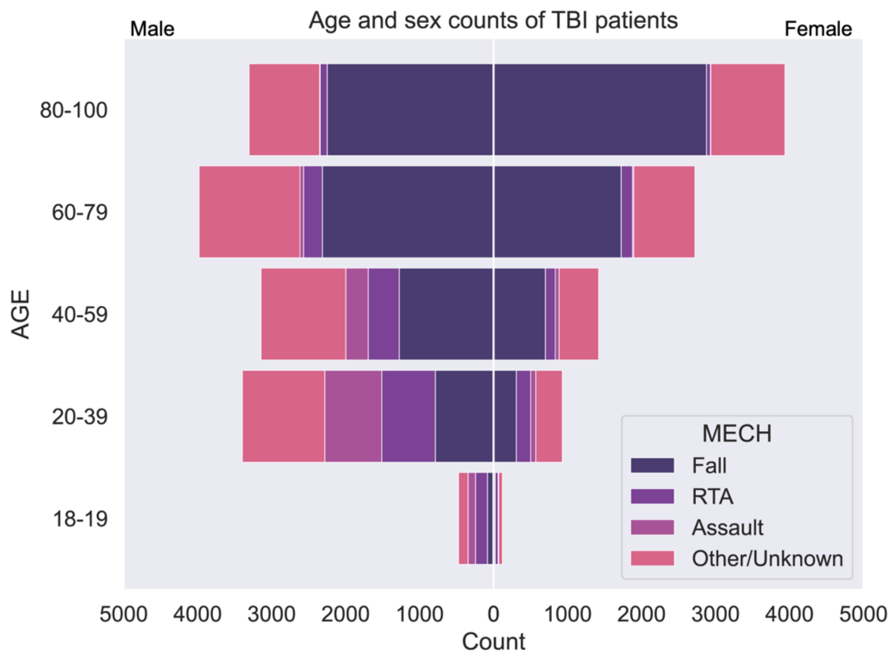

**Supplementary Figure 3. Mechanisms associated with dementia trauma– post-matching.** *Males and females are mirrored, with males on the left and females on the right. Y-axis depicts five-year age bands, with the youngest group (40-44) at the bottom and the oldest group (95-100) at the top; X-axis depicts proportions of trauma episodes that were associated with the following mechanisms: Falls (Indigo), road traffic accidents (RTAs; dark purple), assault (light purple), and all other or unknown causes (pink).*

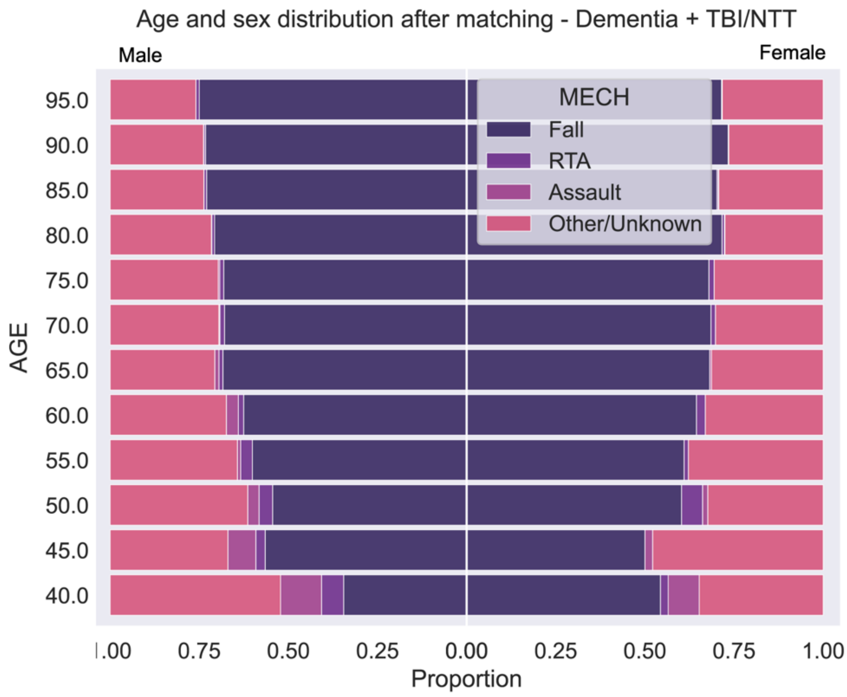

**Supplementary Figure 4. Mechanisms associated with non-dementia trauma – post-matching.** *Males and females are mirrored, with males on the left and females on the right. Y-axis depicts five-year age bands, with the youngest group (40-44) at the bottom and the oldest group (95-100) at the top; X-axis depicts proportions of trauma episodes that were associated with the following mechanisms: Falls (Indigo), road traffic accidents (RTAs; dark purple), assault (light purple), and all other or unknown causes (pink).*

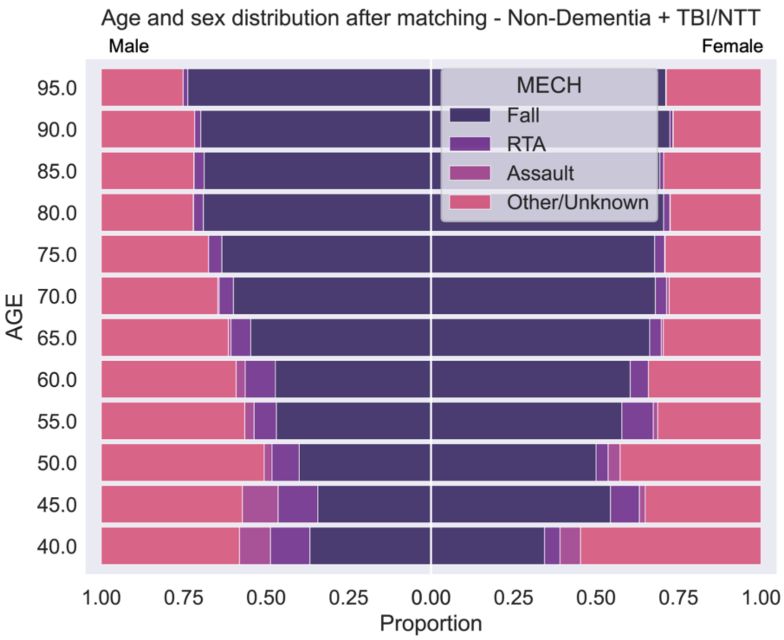

**Supplementary Figure 5. Impact of deprivation on post-injury survival at 1M.** Survival curves depict survival probability at one month after injury (NTT or TBI) stratified by Welsh Index of Multiple Deprivation (WIMD) Quintile in 2019, with quintile 1 representing the most deprived and quintile 5 being the least. X-axis represents time in days and y-axis represents survival probability.

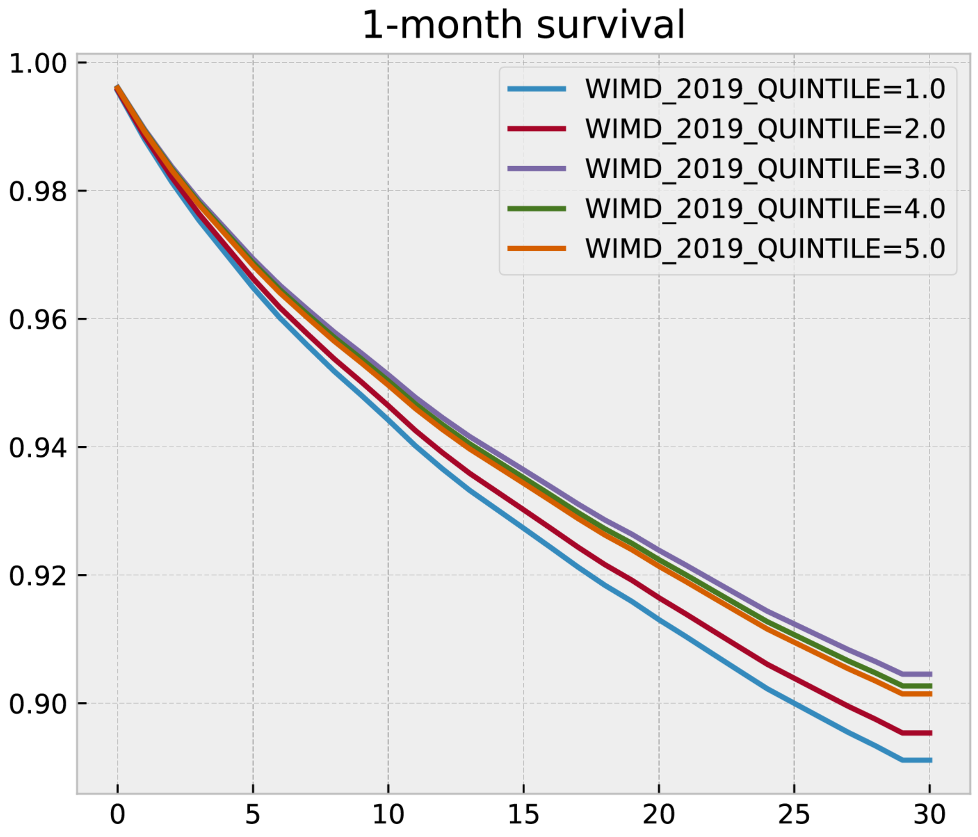

**Supplementary Figure 6. Impact of deprivation on post-injury survival at 6M.** Survival curves depict survival probability at six months after injury (NTT or TBI) stratified by Welsh Index of Multiple Deprivation (WIMD) Quintile in 2019, with quintile 1 representing the most deprived and quintile 5 being the least. X-axis represents time in days and y-axis represents survival probability.

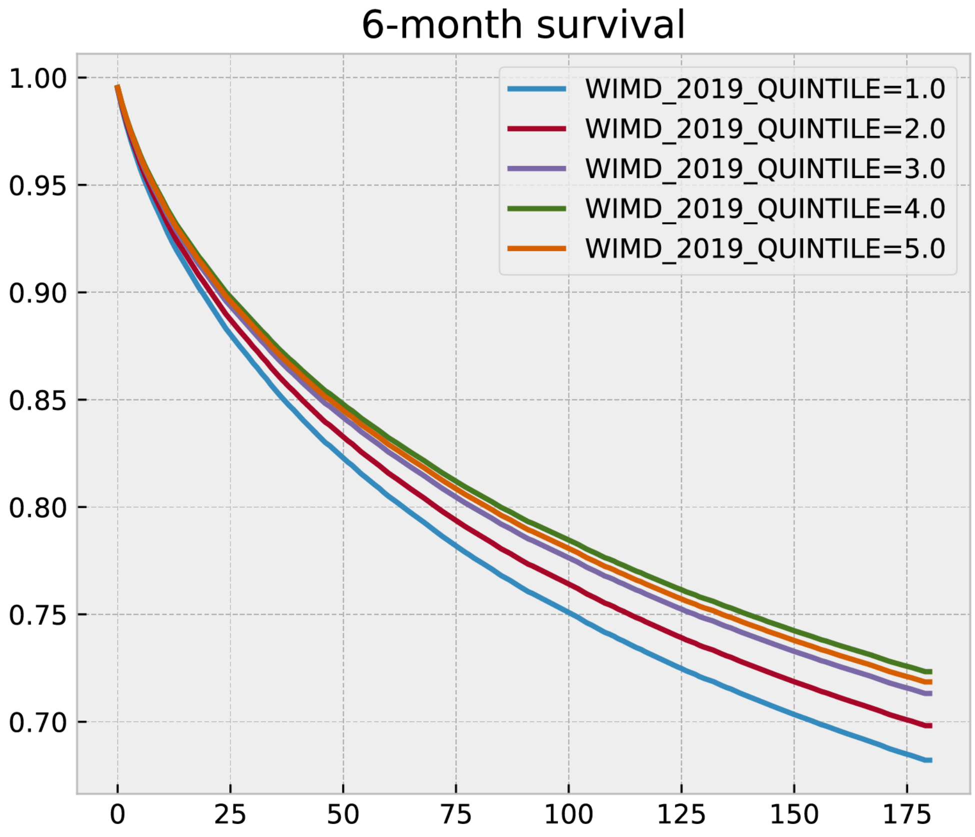

**Supplementary Figure 7. Impact of deprivation on post-injury survival at 12M.** Survival curves depict survival probability at twelve months after injury (NTT or TBI) stratified by Welsh Index of Multiple Deprivation (WIMD) Quintile in 2019, with quintile 1 representing the most deprived and quintile 5 being the least. X-axis represents time in days and y-axis represents survival probability.

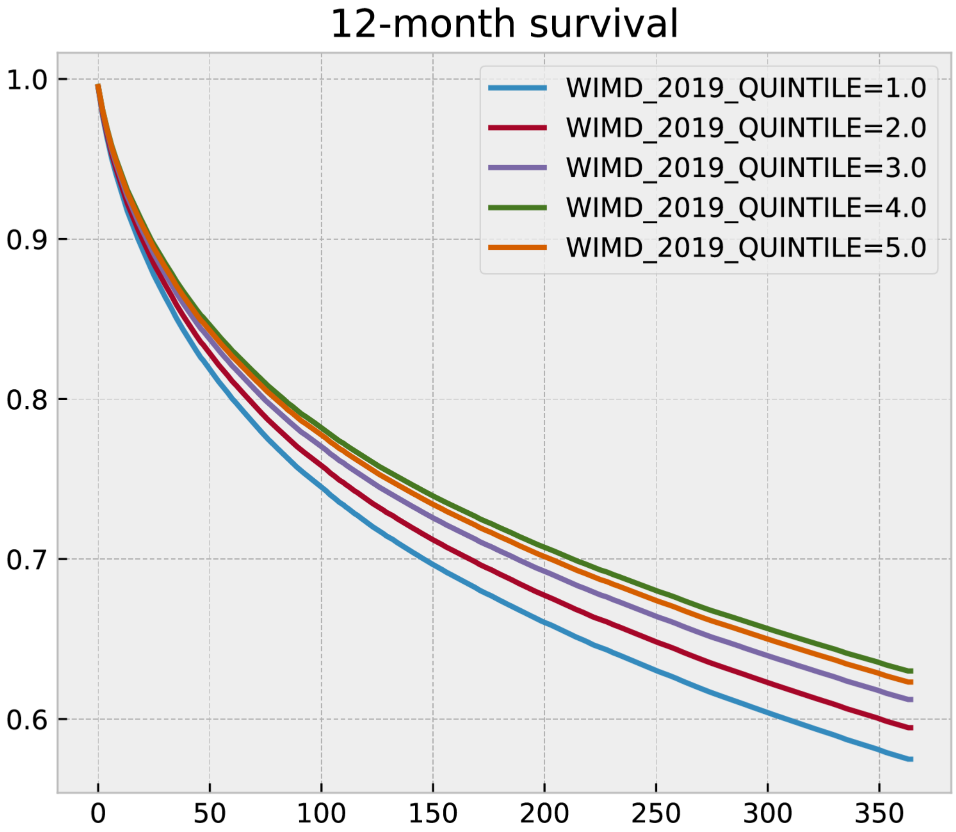

**Supplementary Results 1. Extended cohort description – study population**

Compared to the NTT group, the TBI group were significantly older (TBI mean age=62.6±22.7 years, NTT mean age=59.5±23.9 years, t= -19.1, p<0.001) and contained more men (TBI 61.0% male, NTT 49.8% male, $X^2$ = 1128, p<0.001).

Upon discharge, TBI patients were less likely to be leaving medical care (e.g., returning to own home) (61.8%) compared to NTT patients (80.6%). Rates of ongoing medical care (e.g. transfer to different ward or hospital) were higher in TBI (21.5%) than NTT patients (12.5%), although rates of discharge to social services/ care homes were similar across the groups (3.5% TBI, 3.0% NTT). See Supplementary Table 5 for list of discharge locations and groupings.

**Supplementary Results 2. Extended cohort description – study population with and without dementia**

Among those aged 40 and over, those with dementia were older at the time of injury (DEM+NTT: 83.1±9.3 years; DEM+TBI: 81.5±10.9 years) compared to those without dementia (ND+NTT: 69.4±15.7 years; ND+TBI: 69.9±15.2 years), (One way ANOVA: F(449,043)=14929, p<0.0001) (Table 2). There were significant sex differences ($X^2$ = 4716, p<0.001), with more females with dementia and NTT (DEM+NTT) more likely to be female (69.2%).

**Supplementary Results 3. Extended description of NTT injury mechanisms**

Falls were the most common associated mechanism for NTTs in older age. Falls accounted for 63% of injuries in women and 42% of men aged 55-60, increasing to 80% and 78% respectively in women and men aged 85-100.

**Supplementary Results 4. Extended description of Cox regression results – impact of injury on survival; influence of sex and deprivation**

Male sex was also associated with higher risk of mortality after TBI, compared with females, across all timepoints (1M: 1.45 [1.40,1.50], p<0.0001; 6M:1.42 [1.39,1.45], p<0.0001; 1Y: 1.40 [1.37,1.43], p<0.0001).

Deprivation differentially impacted survival across time. Compared to the most deprived quintile (Q1), the three least deprived quintiles (Q3-Q5) were associated with lower mortality at one month (Q3: 0.87, [0.82,0.92]; Q4: 0.89, [0.84,0.94]; Q5: 0.90, [0.85,0.95]). All quintiles (Q2-Q5) were associated with significantly lower mortality than the most deprived quintile (Q1) at six and twelve months. Mortality was particularly low in the least deprived quintiles (Q4-Q5) (6M: Q2: 0.94, [0.91,0.97]; Q3: 0.88, [0.86,0.91]; Q4: 0.85 [0.82,0.88]; Q5: 0.86 [0.83,0.89]. 12M: Q2: 0.94 [0.91,0.97]; Q3: 0.87 [0.86,0.91]; Q4: 0.83 [0.81,0.86]; Q5: 0.85 [0.83,0.88])

**Supplementary Results 5. Extended description of Cox regression results – impact of injury and dementia on survival; influence of sex and deprivation**

Male sex was associated with higher mortality across the timepoints (1M: 1.33 [1.25,1.41], p<0.0001; 6M: 1.36 [1.31,1.41], p<0.0001; 1Y: 1.37 [1.32,1.41], p<0.0001). For full statistical results, please see Supplementary Tables 16-21.

Deprivation was not associated with significant differences in mortality at one month. At six and twelve months, only Q4 was associated with reduced mortality relative to the most deprived quintile (Q1) (6M: 0.89 [0.84,0.94]; 12M: 0.88 [0.83, 0.93]), with no significant differences in Q2, Q3, and Q5.
